# Population-Scale Precision Safety in Oncology Reveals Clinical and Genetic Determinants of Systemic Therapy Toxicity

**DOI:** 10.64898/2026.09.16.26363259

**Authors:** Ziad Bakouny, X. Alex Guo, Feiyang Huang, Saksham Mohan, Rohan Walser, Zeyun Lu, Tomin Perea-Chamblee, Lamiah Khan, Nishat Ahmed, Antonio Ocejo, A. Ari Hakimi, Neil Shah, Martin H. Voss, Kathryn C. Arbour, Yee-Ming Melody Cheung, Hassan Azhari, David M. Faleck, Rachel Niec, Mark TA Donoghue, John J. Orgera, Aijazuddin Syed, Michael F. Berger, Michele Waters, Karl Pichotta, Christopher Fong, Justin Jee, Nikolaus Schultz, Deborah Schrag, Lonny Yarmus, Adam J. Schoenfeld, Alexander Gusev, Ritesh R. Kotecha, Robert J. Motzer, Craig B. Thompson, Wesley Tansey, Jian Carrot-Zhang, Ed Reznik

**Author notes:** Correspondence to: Z.B., W.T., J.C.Z., E.R. These authors contributed equally to this work (Ziad Bakouny, X. Alex Guo, Feiyang Huang, Saksham Mohan, Rohan Walser). These authors contributed equally to this work (Ziad Bakouny, Wesley Tansey, Jian Carrot-Zhang, Ed Reznik).

## Abstract

Treatment toxicity constrains the use of effective cancer therapies, but its clinical and genetic determinants remain poorly defined. We developed a large language model-based approach to produce MSK-Tox, a pan-cancer resource capturing the incidence, temporality, and grade of toxicity to anti-cancer therapy across more than 50,000 patients. Analysis of six representative adverse events—pneumonitis, adrenal insufficiency, liver toxicity, colitis, hyperthyroidism and hypothyroidism—revealed distinct toxicity landscapes shaped by cancer type, treatment regimen, and clinical context. Pretreatment clinical features enabled individualized prediction of toxicity risk across adverse events, supporting risk assessment before therapy initiation. Beyond clinical predictors, we identified two modes of germline susceptibility to treatment toxicity: an organ-intrinsic mode, in which germline variation confers risk across systemic therapies, exemplified by a regulatory variant near *FOXE1* associated with hypothyroidism; and an immune-mediated mode, confined to immune checkpoint inhibitor-treated patients, in which HLA-DRB1*15 was a major determinant of adrenal insufficiency. Notably, the same allele predisposes to multiple sclerosis in individuals without cancer, indicating that immune checkpoint inhibition unmasks a latent autoimmune predisposition. These findings provide an empirical basis for a new precision safety paradigm for predicting who will be harmed by a therapy on the same principles that guide prediction of therapeutic benefit.

## Introduction

Precision oncology has largely focused on maximizing response by matching patients to therapies based on the molecular features of their tumors. This paradigm has improved treatment selection by identifying patients most likely to benefit from targeted therapy, immunotherapy, and other systemic therapy approaches^1^. On the other hand, systemic therapy toxicity remains a major source of morbidity in cancer care, contributing to treatment interruption or discontinuation, emergency evaluation, hospitalization, and death^2^. For many systemic therapies, and particularly for immune checkpoint inhibitors (ICI), the biological determinants of toxicity or other adverse events remain largely unknown. In the absence of reliable predictors of toxicity, clinicians make treatment decisions without individualized estimates of harm. Precise toxicity prediction would help to optimize treatment exposure and ultimately improving treatment effectiveness.

Gathering evidence of systemic therapy toxicity at scale remains challenging. Although clinical trials typically include detailed toxicity assessments, trial populations are selected, follow-up is often limited, and adverse event patterns may not represent toxicity patterns observed in routine oncology practice^3^. In electronic health records, treatment-related adverse events are recorded in unstructured clinical notes. Although these notes often contain the most clinically relevant information, including symptom onset, diagnostic reasoning, attribution to therapy, management, severity, and resolution, the scale and complexity of free-text clinical documentation have historically limited its use for toxicity discovery. While some cancer centers collect patient-reported symptoms, these systems rely on prespecified questionnaires and active patient participation, limiting coverage and completeness^4,5^.

Here, we developed a large language model-based approach to characterize systemic therapy toxicities at scale in patients with cancer. We then deployed this approach to 2.77 million clinical notes across 55,406 patients with cancer to produce MSK-Tox (The Memorial Sloan Kettering Systemic Therapy Toxicity Dataset), a multimodal, temporally resolved dataset of patient toxicities, augmented with routine lab values, tumor and germline molecular sequencing, and prior treatment exposures. We combined the unprecedented breadth and depth of systemic therapy toxicity annotations with associated rich clinical and genomic data in MSK-Tox to characterize toxicity incidence and outcome associations across cancer types and treatment modalities, develop pretreatment risk models for adverse event onset, and discover germline predictors of hypothyroidism, pneumonitis, and adrenal insufficiency. Most notably, we identified HLA-DRB1*15 as a major determinant of immune checkpoint inhibitor-associated adrenal insufficiency, with evidence of a gene-dosage effect. Together, our findings and the MSK-Tox platform establish a foundation for bringing toxicity research into the era of precision medicine, extending to treatment safety the same molecularly informed paradigm that has transformed oncology efficacy research for more than 15 years.

## Results

### Annotation of systemic therapy toxicities at scale using natural language processing

Toxicities are often recorded in the medical record in free-text notes written by medical oncologists and subspecialists. To ascertain the incidence, severity, and temporal context of adverse events in our cancer patient population, we developed and benchmarked Tox-Annotator, a retrieval-augmented generation–large language model methodology (RAG-LLM). Tox-Annotator segments clinical notes into chunks, embeds them into a vector database using a language model, and retrieves the top-ranked textual chunks using toxicity-specific query templates. Retrieved chunks are batched into 90-day intervals (see **Methods**). Each 90-day interval is then assigned a confidence score of presence of a pre-specified set of toxicities with a customized LLM prompt (**Figure 1a, Figure S1a-S1b, Supplementary Note 1**). Introduction of the RAG substantially reduced computational cost with minimal effect on accuracy, allowing us to deploy our pipeline at institution-scale (**Figure S1c-S1d**, **Supplementary Tables 1 and 2**). We additionally found that an open-weights model, Llama 4 Maverick, achieved similar performance at a similar or lower cost than other models (**Figure S1g-S1h**, **Supplementary Table 3**), suggesting that, within this class of similarly capable models, model choice was not the primary determinant of performance. We therefore proceeded with Llama 4 Maverick for subsequent analyses. We applied this analytical framework to six systemic therapy toxicities that span distinct organ systems, vary widely in prevalence, and that are usually encountered in the context of immune checkpoint inhibitor therapy: pneumonitis, adrenal insufficiency, liver toxicity, colitis, hyperthyroidism, and hypothyroidism. Of note, five of these correspond to a single CTCAE v5.0 term; liver toxicity has no single equivalent and was defined as a composite of the CTCAE v5.0 hepatocellular and cholestatic laboratory terms, which CTCAE grades separately but which we report as one endpoint (see **Methods**).

**Figure 1:**
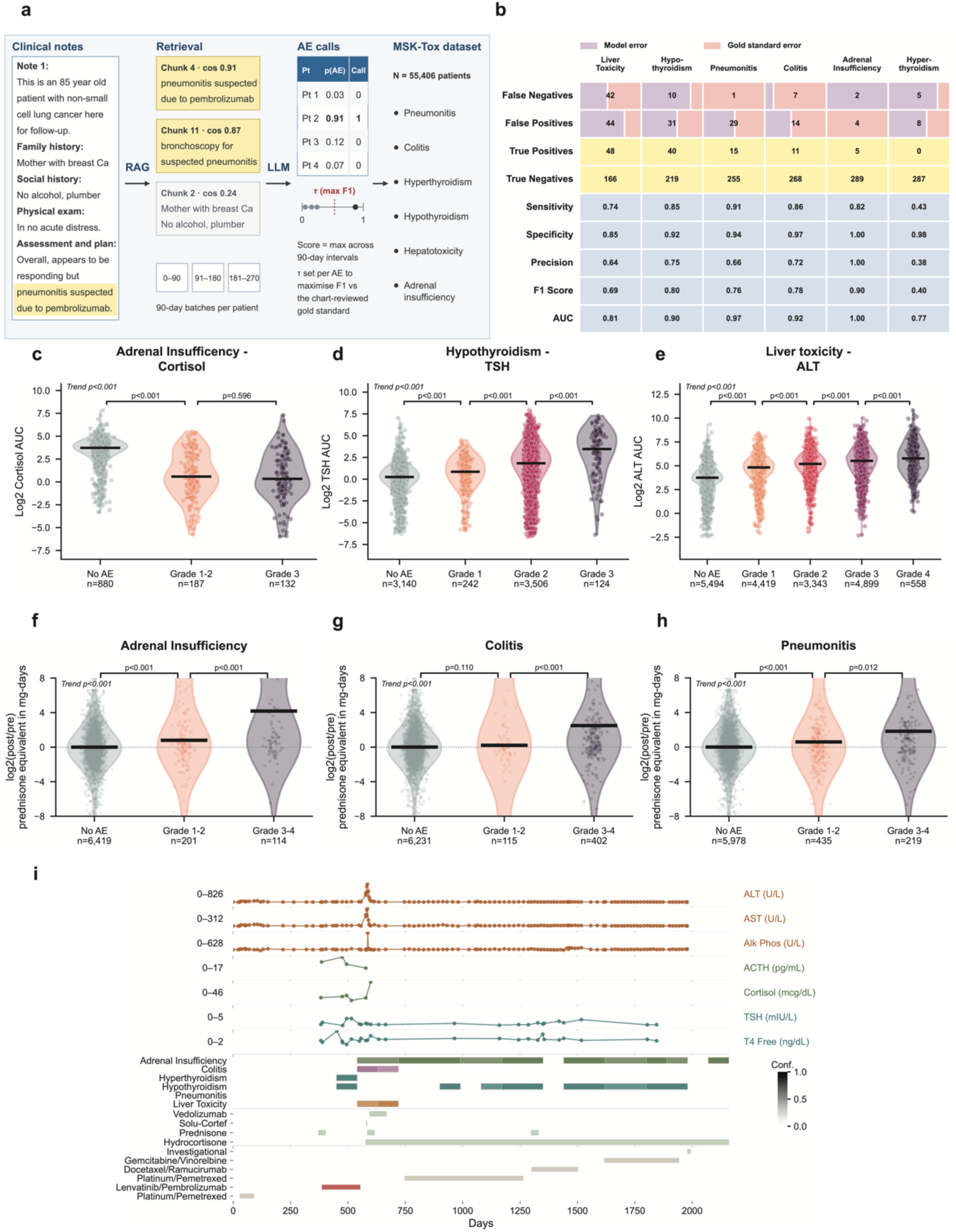
Annotation of systemic therapy toxicities at scale from free text clinical notes using a custom natural language processing pipeline. **a,** Overview of Tox-Annotator for annotating systemic therapy toxicities in MSK-Tox. **b**, Performance metrics for Tox-Annotator against a clinical trial-derived CTCAE-based gold standard in a held-out test set of 300 patients. Orthogonal validation of Tox-Annotator toxicity and grade labels using corresponding laboratory data (**c-e**) and steroid administration data (**f-h**). p-values from Jonckheere-Terpstra trend tests and pairwise Mann-Whitney tests shown. **i**, Example of a timeline of a patient with non-small cell lung cancer treated across multiple lines of systemic therapy with systemic therapy toxicity annotations from Tox-Annotator shown. Top, relevant laboratory data. Second from top, Tox-Annotator annotations for each of the 6 toxicities. Second from bottom, oral and intravenous supportive medications given to treat systemic therapy toxicities shown. Bottom, line of systemic anti-cancer therapy regimens administered.

We took four complimentary approaches to validate the Tox-Annotator predictions, with each source of validation contributing multiple lines of supporting evidence. First, we compared the performance of Tox-Annotator for the 6 adverse events to the gold standard clinical trial-based dataset comprising prospectively collected Common Terminology Criteria for Adverse Events (CTCAE) annotations from 6,279 patients enrolled across 1,057 clinical trials performed between 1/1/2010 and 7/29/2024 at MSK (**Methods**). In a manually reviewed test set of 300 patients, 197 discrepancies (out of 1,800 comparisons) between the Tox-Annotator classification and the clinical trial-based gold standard showed that 44.2% were attributable to errors in the gold standard rather than the Tox-Annotator classification, with most errors being due to only related concepts being reported in the gold standard (*i.e.* dyspnea instead of pneumonitis) or low grade events that are documented but not reported in the gold standard. After adjudication, the pipeline demonstrated strong discrimination across adverse events, with AUCs ranging from 0.77 to 1.00 and F1 scores from 0.40 to 0.90 (**Figure 1b**, **Supplementary Table 4**). Second, we extended this evaluation to a separate validation set of 5,279 patients not included in the initial train/test subset, demonstrating largely consistent, though modestly lower, performance in the absence of manual adjudication of gold-standard discrepancies (**Supplementary Table 5**). The modestly lower performance was likely attributable to uncorrected errors in the clinical trial-based gold standard. Third, we evaluated the consistency of Tox-Annotator-imputed adverse event grade with manually reviewed grade across all 1,264 patients (1,454 comparisons) in the gold standard who had one of the 6 adverse events and had LLM labels. LLM-imputed grade perfectly agreed with gold standard grade in 63.1% of cases, with 28.6% being within 1 level of grade around the gold standard grade (**Figure S1i, Supplementary Table 6,** p < 0.001 versus permutation-based null model). Fourth, we evaluated the performance of Tox-Annotator in accurately determining the time of onset of the AEs compared to the gold standard. To do this, we compared the time of onset of AEs as determined by Tox-Annotator from each line of systemic therapy compared to that in the gold standard and found that Tox-Annotator accurately recapitulated the time of onset of each of the 6 AEs (**Figure S1j-S1k, Supplementary Table 7,** median (LLM – gold standard) = −10.0 days, 95% CI −15.0 to −8.0, Brier score range 0.01 to 0.11, p < 0.001 versus permutation-based null model).

We also sought to validate Tox-Annotator using complementary measurements from the clinical record. To do so, we first evaluated correlations of toxicities with laboratory values and found concordant patterns of laboratory value changes. We observed clinically consistent patterns including decreasing cortisol level with increasing adrenal insufficiency grade (**Figure 1c**, **Supplementary Table 8**, trend test p < 0.001; no AE median log2 (cortisol) [IQR] 3.72 [2.91– 4.26] vs G3+ 0.33 [-0.97–2.42]), increasing TSH with hypothyroidism grade (**Figure 1d**, **Supplementary Table 9**, trend test p < 0.001; no AE 0.25 [-0.67–1.08] vs G3+ 3.46 [1.35–5.11]), and increasing ALT level with liver toxicity grade (**Figure 1e**, **Supplementary Table 10**, trend test p < 0.001; no AE 3.75 [3.19–4.32] vs G4+ 5.77 [4.58–6.90]).

Second, we examined patterns of therapeutic administration following the initiation of adverse events. Typically, patients experiencing certain AEs are treated with systemic corticosteroids to counter autoimmunity or replace endogenous steroid production (*i.e.* in adrenal insufficiency). Consistent with accurate toxicity calls, we observed that cumulative prednisone-equivalent exposure following the imputed toxicity onset (see **Methods**) increased in a grade-dependent manner for adrenal insufficiency (**Figure 1f**, **Supplementary Table 11**, trend test p < 0.001; no AE median log2 (post/pre) [IQR] 0.00 [-2.18–4.22] vs G3–4 4.18 [0.18–12.75]), colitis (**Figure 1g**, **Supplementary Table 12**, trend test p < 0.001; no AE 0.00 [-2.12–3.72] vs G3–4 2.49 [-0.31– 13.61]), and pneumonitis (**Figure 1h**, **Supplementary Table 13**, trend test p < 0.001; no AE 0.00 [-2.18–4.13] vs G3–4 1.83 [-0.52–11.23]). Crucially, although Tox-Annotator–derived toxicity phenotypes correlated with laboratory abnormalities and steroid administration, there remained substantial variation within each toxicity grade, highlighting the difficulty of accurately inferring complex clinical phenotypes from structured data alone. This is consistent with prior studies demonstrating the limitations of laboratory values, medication data, and ICD-9/10 codes when used in isolation to identify treatment-related toxicities^6–8^. Importantly, the Tox-Annotator approach partially addresses this limitation by identifying only toxicities considered attributable to anti-cancer therapy (treatment-related adverse events, TRAE), a distinction that is difficult to make from structured data alone, which could only identify treatment-emergent adverse events (TEAE). For instance, this approach allows us to differentiate treatment-related liver injury from liver enzyme elevations due to other causes, including cancer itself^7^.

In total, these multiple lines of supporting validation establish the accuracy of the Tox-Annotator approach for the identification of AEs from unstructured medical oncology notes. This approach allowed us to quantitatively and temporally define the landscape of systemic therapy AEs across all lines of therapy for patients at MSK (**Figure 1i**).

### Landscape of systemic therapy toxicities

We deployed the above methodology to comprehensively characterize the incidence, severity, and timing of each of the six previously specified AEs across our full cohort of patients who had had molecular testing and were treated with systemic therapy regimens, which included 55,406 patients and 2.77 million medical oncology notes. Demographically, the cohort largely reflected the epidemiology of patients who undergo clinical sequencing as part of routine clinical care at MSK. The median age of patients in our cohort was 63.2 years (IQR 52.9 – 71.4), and 28,847 (52.1%) patients were female. The median follow-up time was 52.0 months (95% CI 51.1 – 52.8). The most common cancer types in our cohort were non-small cell lung cancer (5,831 patients, 10.5%), colorectal cancer (5,164 patients, 9.3%), and breast cancer (4,330 patients, 7.8%). During the first line of systemic therapy treatment, patients were treated with a variety of systemic treatment modalities including chemotherapy alone (29,160 patients, 52.6%), targeted therapy alone (3,347 patients, 6.0%), immune checkpoint inhibition alone (3,145 patients, 5.7%), or combinations of different modalities such as chemotherapy + immune checkpoint inhibition (2,468 patients, 4.5%) and hormone therapy + chemotherapy (1,236 patients, 2.2%) (**Figure S2a**, **Supplementary Table 14**).

In total, during the first line of systemic therapy treatment we identified 11,607 distinct adverse events across the six AEs of interest (liver toxicity, n = 6,191; hypothyroidism, n = 2,760; pneumonitis, n = 1,256; colitis, n = 970; adrenal insufficiency, n = 313; hyperthyroidism, n = 117), including 8,889 Grade 1-2 events and 2,718 Grade 3+ events (**Figure S2b**, **Supplementary Table 15**). Along the temporal clinical trajectory during first-line therapy, the overall proportion of patients encountering these AEs increased with time and differed by toxicity type (**Figure 2a**, **Supplementary Tables 16 and 17**). At 1 year after start of first line of therapy (LoT), liver toxicity had the highest cumulative incidence at 13.3% (95% CI 12.9 −13.7), followed by hypothyroidism at 6.1% (95% CI 5.8 −6.3), pneumonitis at 2.7% (95% CI 2.5 – 2.8), colitis at 2.1% (95% CI 1.9 - 2.2), adrenal insufficiency at 0.7% (0.7 – 0.9), and hyperthyroidism at 0.2% (95% CI 0.2 - 0.3) (**Figure 2b**, **Supplementary Table 18**).

**Figure 2:**
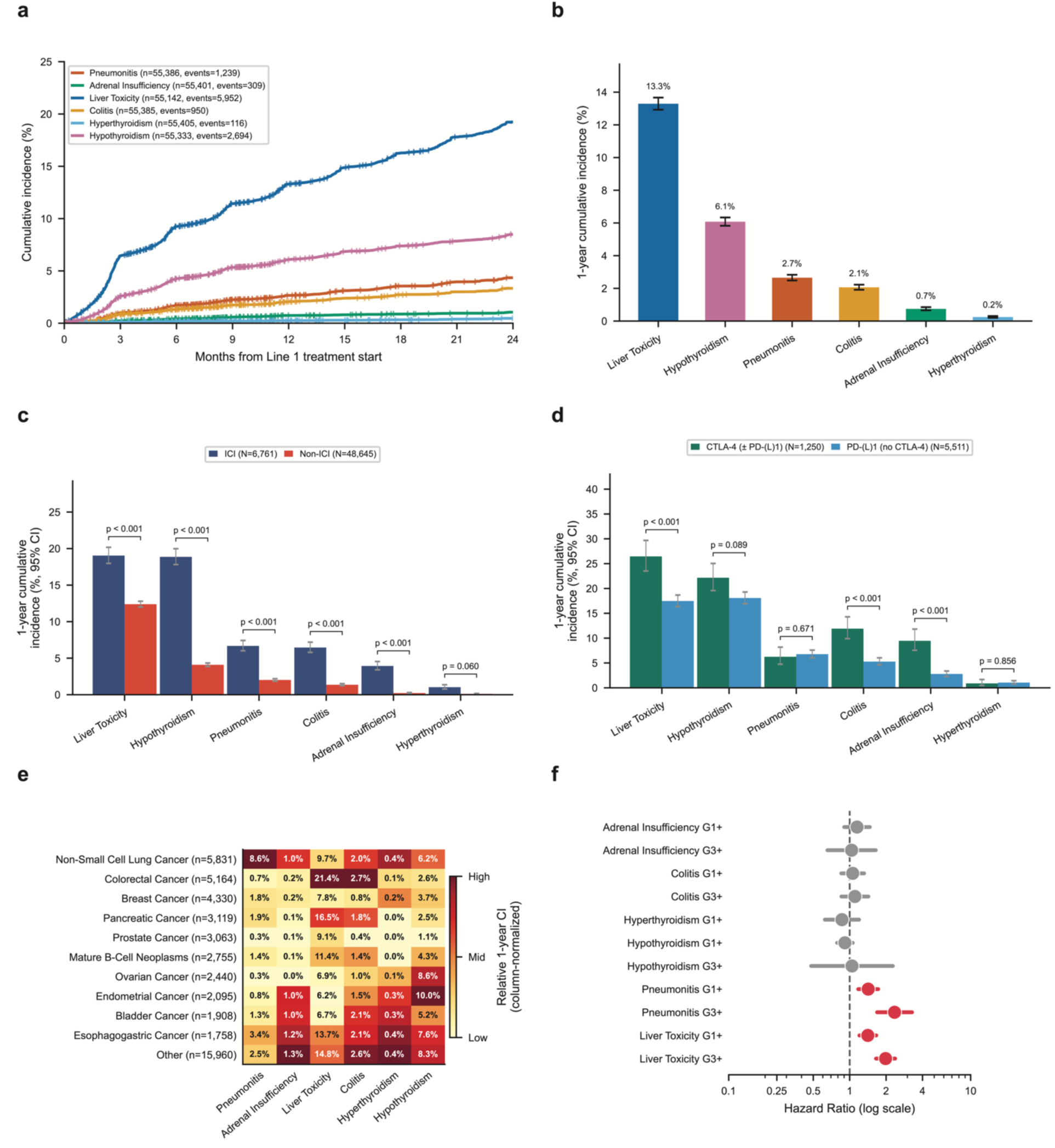
Landscape of systemic therapy toxicities across a pan-cancer cohort. **a,** Cumulative incidence of systemic therapy toxicities in patients treated on first line systemic therapy regimens. **b**, 1-year cumulative incidence and 95% confidence intervals of systemic therapy toxicities on first line systemic therapy regimens**. c,** 1-year cumulative incidence and 95% confidence intervals among patients treated with ICI or non-ICI based regimens. p-values from multivariable cox regression models are shown. **d,** Among ICI-treated patients, 1-year cumulative incidence and 95% confidence intervals among patients treated with CTLA-4-based (+/-PD(L)-1) or PD(L)-1 based regimens. p-values from multivariable cox regression models are shown. **e**, 1-year cumulative incidence of toxicities across different cancer types (including all systemic therapy regimens). The heatmap is colored by column-level normalization. **f**, Among ICI-treated patients, association of adverse event occurrence with progression free survival (NLP-derived) is shown across any grade of toxicity (G1+) and for high grade toxicity alone (G3+). Hazard ratios with 95% confidence from multivariable time-varying cox regression models are shown.

Toxicity patterns differed by sex across systemic therapies even after adjusting for multiple confounders (**Methods**). Women had a lower 1-year cumulative incidence of liver toxicity than men (11.7% [95% CI, 11.2–12.2] vs. 15.0% [14.5–15.6], p<0.001), but a higher incidence of hypothyroidism (7.3% [95% CI, 6.9–7.7] vs. 4.8% [4.5–5.1], p<0.001), with no significant differences in pneumonitis, colitis, adrenal insufficiency, or hyperthyroidism (**Figure S2c**; **Supplementary Table 19**). Age was likewise associated with distinct and directionally consistent toxicity patterns across systemic therapies (**Figure S2d**, **Supplementary Table 20**). For example, rates of liver toxicity decreased progressively with increasing age, with 1-year cumulative incidence declining from 17.3% (95% CI, 16.4–18.3) among patients aged <50 years to 14.3% (95% CI, 13.6–15.0), 10.9% (95% CI, 10.3–11.5), and 8.6% (95% CI, 7.4–10.0) among patients aged 50–65, 65–80, and >80 years, respectively (adjusted HR vs. <50 years: 0.90, 0.78, and 0.72, respectively; all p<0.001). In contrast, hypothyroidism increased from 4.0% (95% CI, 3.6–4.5) among patients aged <50 years to 6.6% (95% CI, 6.1–7.1), 6.8% (95% CI, 6.4–7.3), and 7.8% (95% CI, 6.6–9.2) across progressively older age groups, with significantly increased adjusted risk in the 50–65 and 65–80 groups (both p=0.001). Pneumonitis similarly increased with age, from 1.6% (95% CI, 1.3–1.9) among patients aged <50 years to 3.7% (95% CI, 3.3–4.1) among those aged 65–80 years (adjusted HR 1.12, p=0.002), whereas colitis, adrenal insufficiency, and hyperthyroidism showed no consistent age-dependent pattern (**Figure S2d; Supplementary Table 20**). Prior studies indicate that older age does not uniformly increase anticancer treatment toxicity, with toxicity patterns depending on treatment class and organ system^9,10^. The lower liver toxicity but higher hypothyroidism and pneumonitis rates observed here may therefore reflect a combination of age-related changes in organ and immune physiology and systematic differences in treatment selection and exposure across age groups, rather than a generalized increase in susceptibility to toxicity with aging.

Consistent with expectation, the prevalence of these six toxicity types in MSK-Tox was strongly associated with exposure to ICI therapy. Among patients receiving first-line systemic therapy, ICI-based regimens were associated with significantly higher rates of liver toxicity (1-year cumulative incidence, 19.0% vs. 12.4%; adjusted HR, 1.40), hypothyroidism (18.9% vs. 4.1%; adjusted HR, 2.15), pneumonitis (6.7% vs. 2.0%; adjusted HR, 1.37), colitis (6.5% vs. 1.4%; adjusted HR, 1.43), and adrenal insufficiency (3.9% vs. 0.2%; adjusted HR, 1.31) compared with non-ICI-based regimens. All associations were statistically significant (all p<0.001, **Figure 2c, Supplementary Tables 21 and 22**). Furthermore, among ICI-treated patients, CTLA-4-based regimens were associated with significantly higher rates of liver toxicity (1-year cumulative incidence, 26.5% vs. 17.5%; HR, 1.35), colitis (11.9% vs. 5.3%; HR, 1.46), and adrenal insufficiency (9.5% vs. 2.8%; HR, 1.44) compared with PD-(L)1-based regimens without CTLA-4 blockade. Rates of hypothyroidism were numerically higher with CTLA-4-based regimens but did not differ significantly (22.2% vs. 18.1%; HR, 1.11), whereas rates of pneumonitis and hyperthyroidism were similar between groups (**Figure 2d; Supplementary Tables 23 and 24)**. Notably, the effect of CTLA-4 exposure was organ-specific rather than a uniform increase across adverse events.

Prior pan-cancer studies have suggested substantial heterogeneity in toxicity profiles across malignancies, but have predominantly relied on pooled clinical-trial data or study-level incidence estimates^11–13^, while real-world studies have generally either included substantially smaller populations or relied on administrative definitions of toxicity^14–16^. In MSK-Tox, we observed that adverse event profiles showed prominent organ-specific associations by cancer type (**Figure 2e, Supplementary Table 25**). Across 10 individual cancer types, each represented by >1,700 patients, we identified 19 statistically significant cancer type–toxicity associations in multivariable Cox models (**Methods**). Several of the resulting associations appeared to track the tissue involved by the underlying malignancy. Most notably, non-small cell lung cancer was independently associated with substantially increased pneumonitis risk (HR=1.55 [95% CI, 1.44– 1.67], p<0.001), whereas colorectal cancer was associated with increased liver toxicity (HR=1.53 [1.44–1.62], p<0.001) and colitis (HR=1.10 [1.01–1.20], p=0.036), and pancreatic cancer with increased liver toxicity (HR=1.23 [1.14–1.34], p<0.001). Importantly, these associations persisted after adjustment for treatment class, suggesting that they were not attributable solely to differential use of systemic therapies across malignancies. These findings are consistent with earlier studies suggesting that the site of the primary tumor or metastasis may influence the anatomical pattern of treatment toxicity^14^, while large clinical-trial datasets have also demonstrated cancer-specific differences in adverse event spectra^11,13^. Together, these findings suggest that susceptibility to treatment-related toxicity may reflect an interaction between systemic therapy and the tissue-specific disease context. Potential contributors include pre-existing local tissue injury, metastatic involvement, and prior local therapies (*e.g.* radiation therapy) which could decrease organ-specific reserve and predispose the organ to systemic therapy-induced toxicity^14^.

Finally, we observed that toxicity risk increased across successive lines of systemic therapy, with higher 1-year cumulative incidences of liver toxicity, hypothyroidism, and pneumonitis at line 4+ than in the first line (**Figure S2e; Supplementary Table 26**). These associations persisted in patient-level frailty Cox models accounting for repeated treatment lines and treatment and patient characteristics. Compared with first-line therapy, line 4+ was associated with increased risks of liver toxicity (HR 1.67), hypothyroidism (HR 3.52), pneumonitis (HR 2.03), and adrenal insufficiency (HR 6.96), whereas no significant increase was observed for colitis or hyperthyroidism (**Supplementary Table 27**). The relationship between toxicity and line of therapy has been comparatively understudied, and prior work has largely been restricted to individual treatment classes. To our knowledge, the present analysis is the first to systematically characterize organ-specific toxicity across successive lines of therapy at the patient level across cancer types and systemic treatment classes, while accounting for repeated treatment exposure and treatment composition. Our findings therefore identify a distinct real-world pattern of increasing risk for several clinically important toxicities with cumulative lines of therapy, potentially reflecting cumulative and delayed effects of prior treatment, evolving treatment combinations, and decreasing physiologic reserve in increasingly heavily pretreated patients.

### Toxicity type and grade shape both prognosis and clinical course

Prior studies have reported conflicting associations between immune-related adverse events (irAEs) and outcomes among patients treated with immune checkpoint inhibitors (ICIs). Meta-analyses have generally associated irAEs with improved progression-free survival (PFS) and overall survival (OS), including favorable associations for endocrine and dermatologic toxicities^17,18^. However, studies accounting for the timing of toxicity and immortal-time bias have remained discordant, with some identifying improved outcomes associated with irAEs^19^ and others finding no significant association^20^. To systematically evaluate this relationship, we modeled each toxicity as a time-varying exposure in multivariable Cox models among 6,407 patients receiving first-line ICI-containing regimens, stratified by cancer type and adjusting for demographics and concomitant systemic therapies (see **Methods**). For most of the six toxicities evaluated, there was no significant association with PFS or OS (**Figure 2f, Figure S2f, Supplementary Tables 28 and 29**). In contrast, pneumonitis was associated with shorter PFS (HR 1.42 [95% CI, 1.18–1.72], p<0.001) and OS (HR 1.54 [1.26–1.88], p<0.001), as was liver toxicity (PFS HR 1.41 [1.20–1.66], p<0.001; OS HR 1.59 [1.37–1.84], p<0.001). These associations were stronger for grade ≥3 pneumonitis (PFS HR 2.35 [1.69–3.28], p<0.001; OS HR 2.79 [1.98–3.92], p<0.001) and grade ≥3 liver toxicity (PFS HR 1.98 [1.65–2.38], p<0.001; OS HR 2.64 [2.18–3.18], p<0.001). Grade ≥3 colitis was additionally associated with shorter OS (HR 1.32 [1.01–1.72], p=0.043). Higher-grade toxicity was also associated with worse outcomes among patients receiving non-ICI therapy (**Supplementary Tables 28 and 29**), suggesting that the adverse prognostic effect of severe toxicity may reflect treatment interruption or discontinuation, morbidity, and subsequent cancer progression rather than an ICI-specific link between immune activation and antitumor efficacy. Importantly, this analysis was restricted to the six toxicities captured by our pipeline (liver toxicity, pneumonitis, colitis, adrenal insufficiency, hypothyroidism, and hyperthyroidism) and should not be generalized to all toxicities. Other toxicities, particularly dermatologic and additional endocrine toxicities, were not evaluated and have been associated with favorable outcomes in prior studies among ICI-treated patients^18,21^. Overall, with conclusions restricted to the six toxicities under study here, our results argue against a uniform association between irAEs and improved ICI efficacy and instead demonstrate that outcome associations depend strongly on toxicity type and severity.

The clinical course of treatment-related toxicities is heterogeneous, and understanding whether severe events arise abruptly or evolve from preceding lower-grade toxicity may inform monitoring and early intervention. Prior longitudinal studies have characterized time to onset, resolution, chronicity, and progression of lower-grade toxicities to grade ≥3 events, demonstrating substantial differences across organ systems^22–24^. However, these studies have generally focused on toxicity onset and resolution rather than reconstructing the within-patient trajectory surrounding a severe event. We therefore characterized toxicity grade at 90-and 180-day intervals before and after the peak grade ≥3 event across patients receiving systemic therapy (**Figure S2g**, **Supplementary Table 30**).

Distinct organ-specific trajectories emerged. Hypothyroidism showed substantial antecedent and persistent lower-grade toxicity, present in 67% and 83% of patients at 180 and 90 days before the grade ≥3 event and in 92% and 83% at 90 and 180 days afterward. Adrenal insufficiency similarly remained present in 88% and 83% of patients at 90 and 180 days after the severe event. In contrast, colitis was largely abrupt and resolved rapidly, with lower-grade toxicity present in only 6% and 8% of patients at 180 and 90 days before the grade ≥3 event and 14% and 8% at 90 and 180 days afterward. Liver toxicity frequently evolved from lower-grade abnormalities, present in 61% and 64% of patients before and 64% and 59% after the severe event, whereas pneumonitis showed an intermediate pattern, with lower-grade toxicity present in 25%–26% before and 58% and 42% after the event. This reveals that severe toxicity is not a uniform binary event, but rather that some grade ≥3 toxicities represent escalation of an already detectable lower-grade process, potentially providing a window for closer surveillance, whereas others emerge more abruptly and resolve more rapidly.

Overall, our analyses establish that systemic therapy toxicities display marked heterogeneity in their incidence rates, timing, and severity across treatments and cancer types. The enrichment of specific toxicities within therapeutic and disease context suggests that adverse event risk is shaped jointly by treatment exposure and host or tumor factors, rather than by therapy class alone. Our results highlight the need for toxicity-specific and context-aware risk stratification and early interventions aimed at preserving treatment continuity and improving clinical outcomes.

### Machine learning models stratify risk of systemic therapy toxicities with clinical features

Numerous tools estimate the efficacy of systemic therapy, ranging from molecular predictive biomarkers^25,26^ to models built from routine clinical and laboratory data^27,28^. Comparatively few such tools exist for toxicity, and risk is instead approximated from incidence within trial arms, in populations narrowed by eligibility criteria^29^. To enable treatment decision support, we developed models of adverse event risk using data available before the decision to treat. We trained random survival forest (RSF) models using patient demographic and clinical features – including current age, sex, cancer type, number of prior therapy lines for each treatment modality, and the planned treatment modality itself. Models were trained to predict time to adverse event from the start of treatment using an 80/20 train-test split of the MSK-Tox dataset, balanced by cancer type and treatment modality (see **Methods, Figure 3a**).

**Figure 3:**
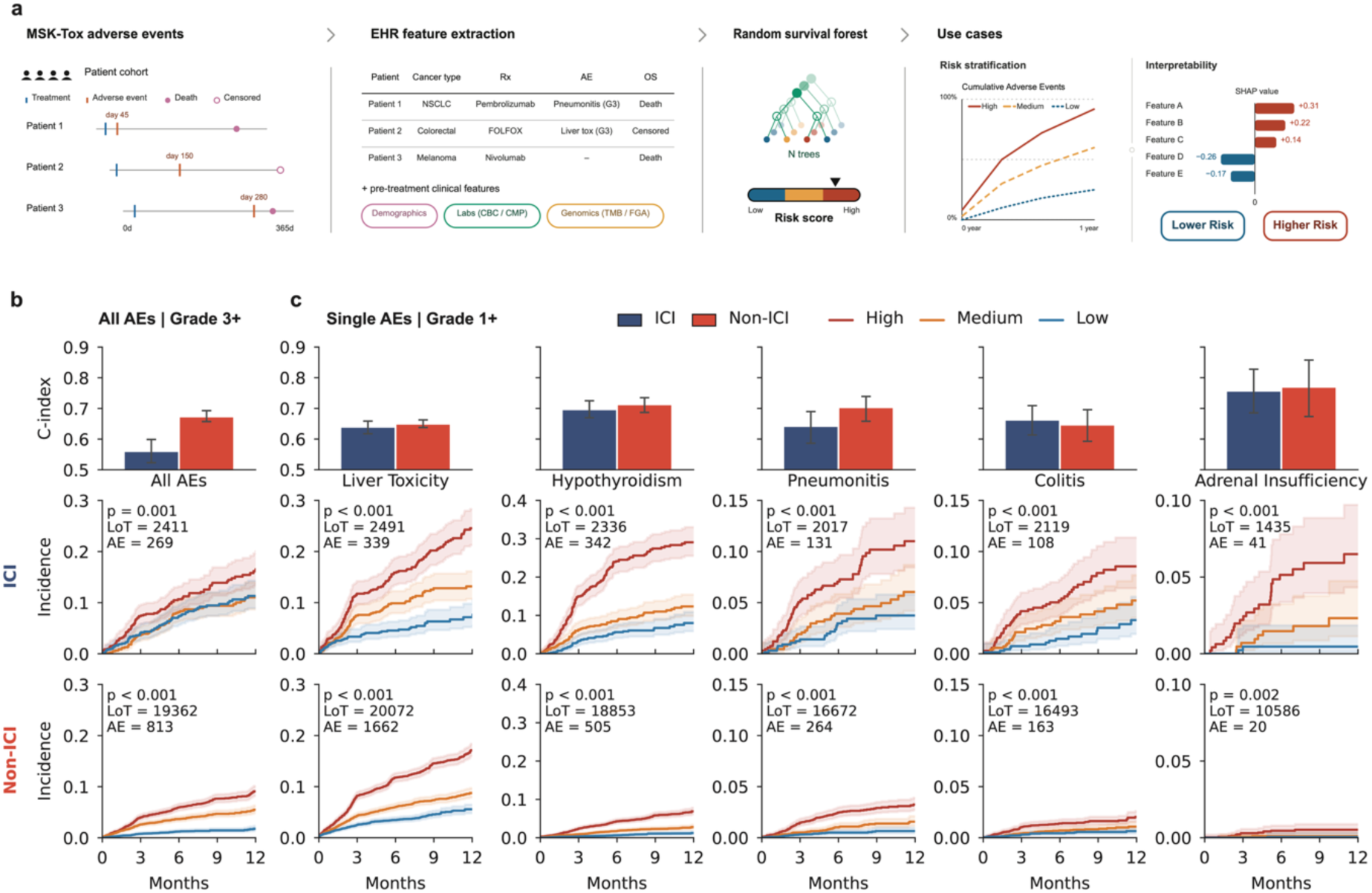
Machine learning models stratify risk of systemic therapy toxicities with clinical features**. a,** Development of random survival forest risk model. **b,** Performance of risk model trained on grade 3+ severe adverse events. **c**, Performance of risk models trained separately on grade 1+ incidence of each adverse event. Harrell’s c-index was calculated on held out test set, and 95% confidence interval was derived from sample bootstrap. 12-month cumulative incidence were shown from the start date of treatment, stratified by ICI and non-ICI regimens, with log-rank p-values.

We first trained a toxicity-type-agnostic model which predicted the risk of onset of any of the six pre-specified adverse events. We measured risk model performance using the Harrell’s concordance index (c-index) for time-to-event prediction. The toxicity-agnostic risk models stratified adverse event risk for both ICI and non-ICI treated patients with modest accuracy. In the grade 1+ setting, the model performed at a c-index of 0.615 (0.591 −0.635) for ICI-treated patients and 0.632 (0.622 - 0.643) for non-ICI treated patients. In the grade 3+ setting, the model performed at a c-index of 0.561 (0.523 - 0.599) for ICI-treated patients and 0.674 (0.657 - 0.693) for non-ICI treated patients (**Figure 3b**, **Supplementary Tables 31–34**).

The modest performance of the toxicity-agnostic model, and the heterogeneity of AE incidence across the clinical cohort and treatment modalities (**Figure 2**) suggest that aggregating heterogeneous adverse events into a single pan-toxicity endpoint could possibly obscure toxicity-specific risk architecture, motivating adverse event–specific prediction models. We therefore trained models to predict risk of a single adverse event type (i.e. colitis only) for each of the six AE classes. We trained AE-specific models in the grade 1+ setting, to ensure sufficient events for model training and stable c-index estimation in the held-out test set. We found that model risk score predicted clinically meaningful differences in absolute risk of developing each specific toxicity surpassing those of AE-agnostic models (**Figure 3c**, **Supplementary Table 35**, hyperthyroidism excluded due to low sample size). For ICI treatments, c-index ranged from 0.640 (0.617 - 0.659) for liver toxicity to 0.758 (0.685 - 0.828) for adrenal insufficiency, and for non-ICI treatments from 0.650 (0.638 - 0.662) to 0.770 (0.673 - 0.858) for the same two toxicities. Stratification by RSF model risk tertiles consistently identified a high-risk population with significantly increased 1-year incidence across all adverse events, both within ICI treated and non-ICI treated groups (**Figure 3c**, **Supplementary Table 36**). In ICI-treated patients for instance, incidence almost tripled in the high-risk group versus the low-risk group for liver toxicity (24.6%; 21.2% - 28.3% vs. 7.1%; 5.2% - 9.8%) and hypothyroidism (29.1%; 25.5% - 33.0% vs. 8.0%; 5.9% - 10.9%), and nearly tripled for pneumonitis (11.0%; 8.4% - 14.3% vs. 3.7%; 2.4% - 5.8%). Model accuracies were largely consistent across lines of therapy (**Figure S3a**) and treatment modalities (**Figure S3b**, **Supplementary Tables 37–43**). Training AE-specific risk models has the potential to capture AE-specific etiologies and risk factors and may be advantageous in deployment, since each AE type would be of particular concern in different clinical contexts or phases of care (e.g. prophylactic treatment, hospitalization).

We next examined the features driving risk prediction in the toxicity-specific RSF models using SHAP attribution. We focused on renal cell carcinoma (RCC) as an illustrative context, as it is one of the few malignancies in which PD-(L)1 blockade, CTLA-4 blockade, and VEGFR-targeted therapy are all encountered in routine practice. Feature attributions were generally concordant with clinically plausible risk factors, and the dominant contributors differed markedly between adverse events.

For liver toxicity, the largest positive contributions came from treatment modality, with CTLA-4 blockade, PD-(L)1 blockade, and targeted therapy each shifting predicted risk upward (**Figure S3c**). This recapitulates our own cohort-level estimates (**Figure 2c–d**) as well as established pharmacovigilance, in which hepatitis is more common with combined CTLA-4/PD-1 blockade than with PD-1 blockade alone^30–32^. The contribution of targeted therapy is expected in this disease, where transaminase elevation is a common toxicity of anti-angiogenic tyrosine kinase inhibitors^33^. Male sex and younger age made smaller but directionally coherent positive contributions, matching the cohort-wide associations we observed across systemic therapies (**Figure S2c–d**).

The attribution profile for hypothyroidism was distinct (**Figure S3d**). PD-(L)1 blockade was the dominant contributor, consistent with thyroid dysfunction being the most common endocrine irAE of PD-1 blockade^34^ and with the increased risk we observed with ICI exposure (**Figure 2c**). Targeted therapy was another strong positive contributor, in keeping with the independent thyrotoxicity of VEGFR-directed tyrosine kinase inhibitors in RCC^35,36^. Male sex was among the strongest negative contributors, consistent with the marked female predominance of autoimmune thyroid disease in the general population^37^ and with meta-analysis of ICI-treated patients associating female sex with ICI-related thyroid dysfunction^38^.

Prior work has suggested that routine lab values might be useful prognostic variables for adverse event risk^39,40^, while the correlation between tumor genomic features and adverse events remains an active debate^41,42^. We therefore studied the predictive value of pre-treatment lab values and tumor genomics in predicting subsequent adverse event risk by training models with additional features derived from routine hematologic and blood lab panels and somatic tumor genomics derived from MSK-IMPACT targeted sequencing (including summary measures such as tumor mutation burden, microsatellite instability, and fraction genome altered). We found that the addition of laboratory variables and/or somatic tumor genomic features did not significantly improve predictive accuracy across adverse events (**Figure S3e**), except for liver toxicity, where the inclusion of lab values led to a small but significant improvement in prognostic accuracy.

Altogether, these findings demonstrate that pre-treatment clinical information alone can stratify patients by risk of developing systemic therapy toxicities. This approach for personalized quantifiable risk prediction could be used in clinical practice, complementary to existing tools for efficacy prediction, to personalize optimal treatment regimen based on a balance of efficacy and toxicity risk.

### Genome-wide association studies identify germline predictors of systemic therapy toxicities

The RSF models above demonstrate that inter-individual variability in cancer-associated adverse events and treatment toxicities can be partly explained by clinical factors, tumor type, and treatment exposure. Previous work has suggested that germline genetic variation, which captures latent, patient-intrinsic differences in immunity and other biological processes, may explain aspects of AE risk^43–46^. However, a systematic investigation of how germline factors shape AE risk is lacking.

To address this, we carried out genome-wide association analysis (GWAS) of AE risk. Building on previous work^43^, we used GLIMPSE to impute germline information from targeted panel sequencing of matched tumor and normal blood from 35,669 patients treated with systemic therapy regimens within the MSK-Tox dataset (including 9,760 ICI treated across any line of therapy, see **Methods**). For these genome-wide association analyses, genotypes restricted to common variants (MAF ≥ 0.01), which were imputed from panel sequencing, showed a mean correlation of 0.88 when compared with true-called genotypes from 88 patients with matching whole-genome sequencing.

Using the MSK-Tox dataset, we carried out a time-to-event GWAS associating the risk of any of the six AEs with germline variation. Because toxicity risk may be influenced by treatment context, we tested for germline associations for any of the six pre-specified toxicities separately in ICI– treated patients, non–ICI–treated patients, and the combined cohort. Interestingly, in both the full cohort of patients as well as sub-cohorts restricted to those patients receiving ICI or non-ICI therapies, we identified no SNPs reaching genome-wide significance for association with risk of pan-toxicity AE (**Supplementary Table 44**). This finding, consistent with our experience in predicting AE risk using RSF models, suggested putative germline factors dictating risk could be specific to one AE, rather than conferring generalized risk to any AE.

Consequently, we carried out a time-to-event GWAS associating the risk of each of the six AEs with the presence of common germline variants. In 3 of 6 AEs (pneumonitis, hypothyroidism, and adrenal insufficiency), we found at least 1 locus achieving genome-wide significance (p<5*10^-8^). To independently validate these findings, we carried out time-to-event GWAS analysis in an orthogonal cohort of 6,112 patients from Dana-Farber Cancer Institute (DFCI) treated with ICI and with similarly imputed genotypes. We additionally evaluated each significant GWAS locus in two population-based resources, FinnGen and UK Biobank; although these cohorts are drawn from the general population and do not capture systemic therapy–related toxicities, we used them to test whether each implicated locus is associated with the corresponding organ-specific disease phenotype in the general population (see **Methods**).

In hypothyroidism, we identified a genome-wide significant association (**Figure 4a**, **Figure S4a**, **Supplementary Table 45**) localized to a regulatory region near *FOXE1* (rs10983761; 9q22.33; MSK-Tox SPACox p= 3.9*10^-10^), a thyroid-lineage transcription factor with established roles in thyroid development, differentiated thyroid function, and adult thyroid homeostasis^47–49^. Importantly, this polymorphism was associated with hypothyroidism across both ICI-and non-ICI-treated patients (**Figure 4a-b**), suggesting modulation of thyroid susceptibility rather than an ICI-specific mechanism. The alternate allele was associated with increased *FOXE1* expression and increased hypothyroidism risk (**Figure S4b**, **Supplementary Table 46**), consistent with prior EMR-linked GWAS showing that common variants near *FOXE1* are associated with primary hypothyroidism and other thyroid phenotypes in patients without cancer^50^. The hypothyroidism risk allele was common in 1000 Genomes European-ancestry reference samples, with an allelic frequency of 66.3% (**Figure S4c**, **Supplementary Table 47**). The association was independently validated in the DFCI cohort (**Supplementary Table 48**), where the lead variant was directly assessable (HR 1.15, 95% CI 1.05-1.26, p = 3.2 × 10⁻³ for endocrine disorders; HR 1.15, 95% CI 1.03-1.28, p = 1.6 × 10⁻² for hypothyroidism), closely matching the discovery estimate in MSK-Tox (HR 1.25, 95% CI 1.13-1.39, **Figure 4b, Supplementary Table 49).** Specifically, this allele reached genome-wide significance for thyroid disease in FinnGen (disorders of the thyroid gland, p = 4×10⁻²⁶⁸; autoimmune hypothyroidism, p = 4×10⁻²⁵¹, **Figure S4d, Supplementary Table 50)** and UK Biobank **(Supplementary Table 51;** hypothyroidism, p = 5×10^-1^^46^). Thus, exposure to systemic cancer therapy unmasks a latent risk for hypothyroidism/thyroid dysfunction.

**Figure 4:**
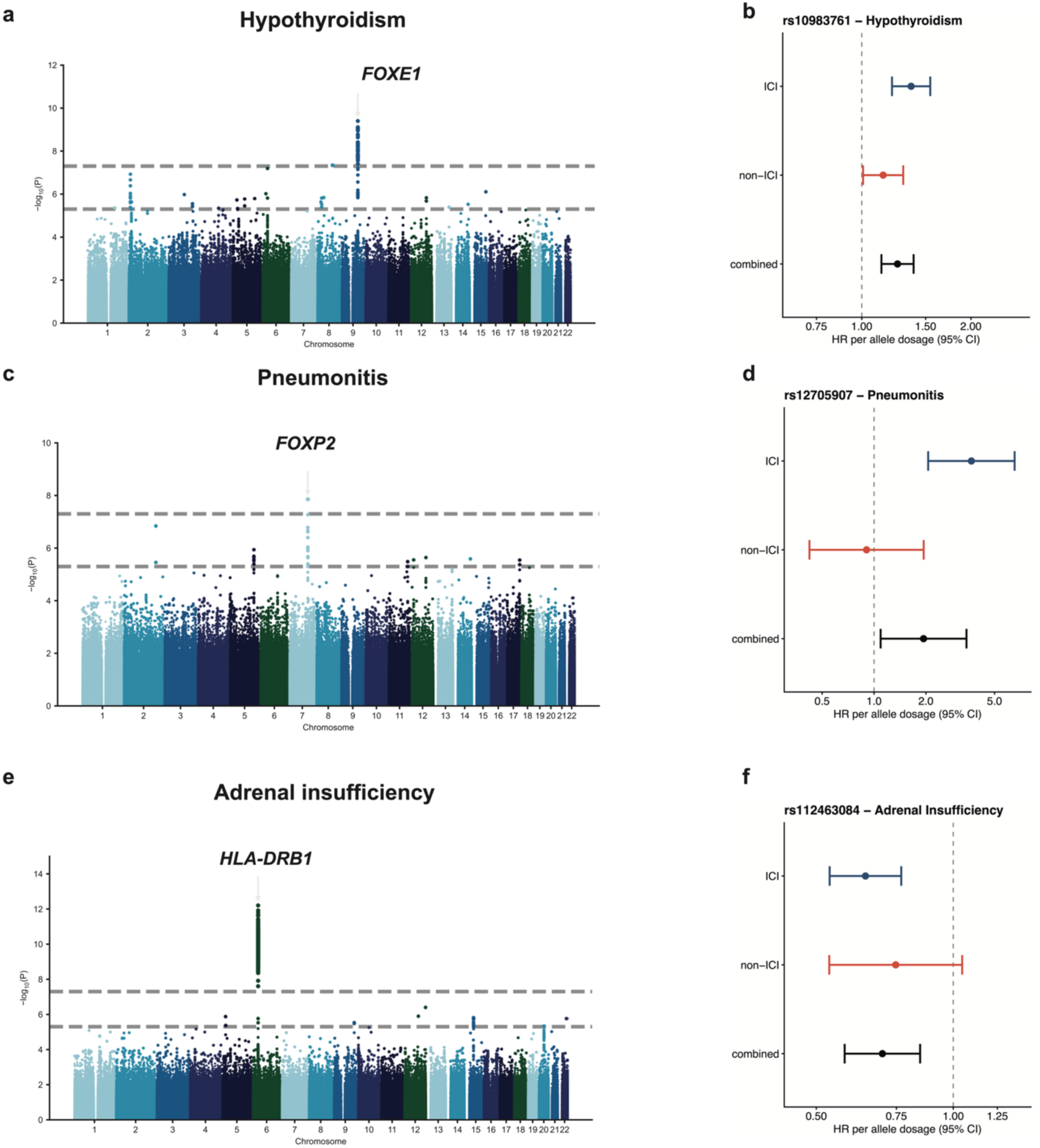
Genome-wide association studies identify germline predictors of systemic therapy toxicities**. a, c, e**, Manhattan plots from multivariable time-to-event genome-wide association studies of hypothyroidism (**a**, combined cohort; 35,669 patients, 1,765 events), pneumonitis (**c**, ICI cohort; 9,723 patients, 661 events), and adrenal insufficiency (**e**, ICI cohort; 9,760 patients, 258 events). P-values were derived from SPACox. Horizontal dashed lines denote thresholds for suggestive (P < 1 × 10⁻⁵) and genome-wide significance (P < 5 × 10⁻⁸). **b, d, f**, Forest plots showing the association between allele dosage of the lead variant from each genome-wide association study and its corresponding phenotype: rs10983761 (*FOXE1*) with hypothyroidism (**b**), rs12705907 (*FOXP2*) with pneumonitis (**d**), and rs112463084 (*HLA-DRB1*) with adrenal insufficiency (**f**). Subdistribution hazard ratios per allele dosage and 95% confidence intervals were estimated using multivariable Fine–Gray models with death as a competing risk, fitted separately within ICI-treated, non-ICI-treated and combined cohorts and adjusted for age, sex and line of therapy. Hazard ratios are plotted on a log scale and the dashed vertical line marks the null. All genome-wide association analyses were run separately for each adverse event on grade 1+ incidence, modelling time to event onset with SPACox and adjusting for sex, age, line of therapy, treatment class, cancer type, sequencing panel and the first ten genetic principal components.

For pneumonitis, we identified a genome-wide significant association (**Figure 4c, Figure S4e**) at rs12705907 (7q31.1; MSKCC SPACox p= 1.4*10^-8^), an eQTL for *FOXP2* (**Figure S4f**). *FOXP2* is a member of the FOXP transcription factor family; *FOXP2* is expressed in respiratory epithelium during lung development, while related FOXP factors regulate lung epithelial injury responses and regeneration^51–53^. The pneumonitis risk allele was less frequent with an allelic frequency of 2.1% and was associated with increased risk of ICI-associated pneumonitis (HR 3.67, 95% CI 2.06-6.54 in the ICI cohort; **Figure 4d**), implicating inherited variation in lung developmental and repair programs as a determinant of immune-mediated pulmonary toxicity. The risk allele was not well imputed in the DFCI cohort and could not be replicated. FOXP2 is expressed in the developing lung epithelium, particularly distal/alveolar epithelium, and FOXP2 cooperates with FOXP1 during lung development^51,54^. In human genetic and functional studies, variants associated with the *FOXP1* gene (closely related to *FOXP2*) have been associated with lung function and idiopathic pulmonary fibrosis risk^55^. Thus, ICI exposure may unmask an organ-intrinsic susceptibility to pneumonitis, in which inherited variation in lung epithelial developmental and repair pathways modulates vulnerability to immune-mediated pulmonary injury.

The strongest GWAS association was for risk of adrenal insufficiency, where a genome-wide significant variant (**Figure 4e, Figure S4g**) in the *HLA-DRB1* region (rs112463084; 6p21.32; MSKCC SPACox p= 6.1*10^-13^) strongly associated with risk of adrenal insufficiency, largely in an ICI-specific manner (**Figure 4f**). The HLA-DRB1 association replicated in the DFCI cohort (best LD proxy rs181159261, r² = 0.81; HR 0.60, 95% CI 0.47-0.77, p = 4.3 × 10⁻⁵ for adrenal insufficiency; and rs9269910, r² = 0.80; HR 0.40, 95% CI 0.26-0.62, p = 3.5 × 10⁻⁵ for severe adrenal insufficiency), closely matching the discovery estimate in MSK-Tox (HR 0.64, 95% CI 0.53-0.77 in the ICI cohort; **Figure 4f**). These data indicate that, in the case of adrenal insufficiency, exposure to ICI, but not other systemic therapy, unmasks a latent risk for autoimmune disease. In contrast to hypothyroidism and pneumonitis, the association between germline variation and risk for adrenal insufficiency was related to a systemic immune-related gene rather than an organ-intrinsic vulnerability. The lead variant was not present in either FinnGen or UK BioBank and therefore could not be assessed for related phenotypes in either dataset.

Finally, a prior study had reported three loci associated with pooled immune-related events^43^. We therefore tested whether any of these loci or their SNP proxies associated with risk across any of the adverse events we tested (combined analysis) and found no association (**Supplementary Table 52**). Instead, in exploratory analyses of individual adverse events among ICI treated patients, proxies of two of the three loci showed nominal, directionally consistent associations; rs16906115 with grade 2+ pneumonitis (best proxy rs16906090, r² = 0.77; p = 5.7 × 10⁻³) and rs113861051 with grade 2+ hypothyroidism (best proxy rs4859281, r² = 0.87; p = 0.014). Both associations were absent in non-ICI-treated patients. This suggests that even while prior studies had found associations with any immune-related adverse events, these results may still have been potentially driven by toxicity-specific associations since hypothyroidism and pneumonitis are among the most frequent irAEs.

Together, the results above shift the germline paradigm for systemic therapy toxicity from a search for risk alleles agnostically associated with immunotherapy-induced adverse events toward a model in which inherited susceptibility is conditional on both the affected organ and therapeutic exposure. Some loci appeared to reflect organ-intrinsic vulnerability, including *FOXE1* for thyroid dysfunction, whereas others pointed to immune-mediated mechanisms of tissue injury. The strongest example was the adrenal insufficiency signal at *HLA-DRB1*, which was restricted to ICI-treated patients. We therefore sought to resolve this association beyond the regional GWAS signal by fine-mapping HLA-DRB1 alleles and antigen-recognition domain residues that might define susceptibility to ICI-associated adrenal insufficiency.

### HLA fine-mapping identifies DRB1*15 as a genetic determinant of ICI-associated adrenal insufficiency

In the genome-wide association study for ICI-related adrenal insufficiency, the strongest association signal localized predominantly to the *HLA-DRB1* locus (chromosome 6:29,649,000-33,368,000 bp), with the lead variants exceeding genome-wide significance (p < 5×10^-8^; **Figure 5a, Supplementary Table 53)**. *HLA-DRB1* is one of the most reproducibly implicated immune-regulatory loci in human autoimmunity, with specific alleles shaping antigen presentation and disease susceptibility^56^, but its role in cancer-therapy-associated toxicities is unclear. Importantly, unlike *FOXE1* and hypothyroidism, the association between variation in *HLA-DRB1* and adrenal insufficiency was specific to patients treated with ICI, with no SNPs achieving genome-wide significance for adrenal insufficiency in the non-ICI setting.

**Figure 5:**
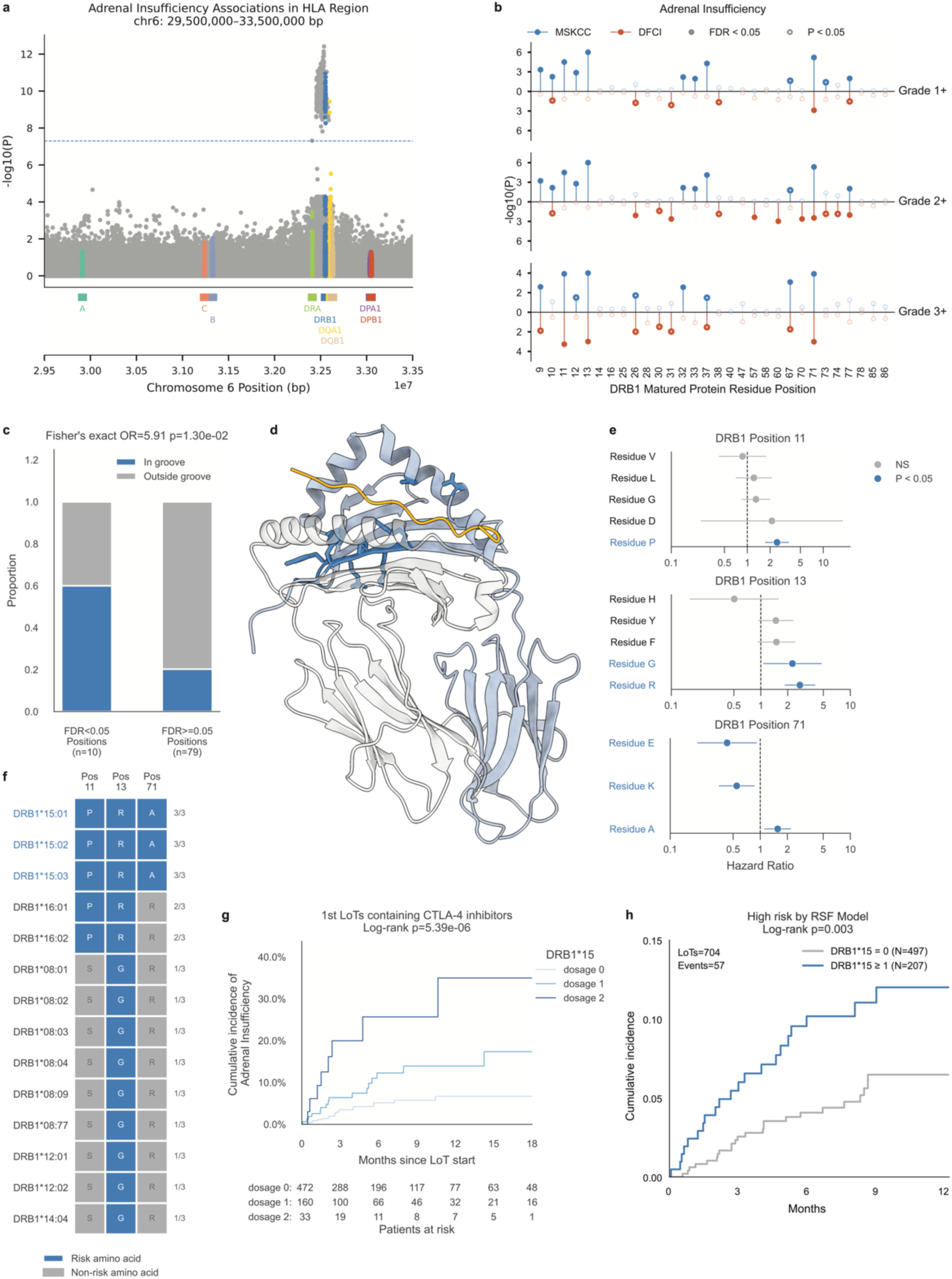
HLA fine-mapping identifies HLA-DRB1*15 as a genetic determinant of ICI-associated adrenal insufficiency. **a,** Regional association plot for adrenal insufficiency across the major histocompatibility complex on chromosome 6 (29.5-33.5 Mb). Variants mapped to classical HLA genes are highlighted by color; the dashed horizontal line denotes the genome-wide significance threshold (P = 5 × 10^-8^). **b,** Amino-acid-level associations across mature HLA-DRB1 protein positions for grade 1+, grade 2+ and grade 3+ adrenal insufficiency in the MSKCC and DFCI cohorts. MSKCC (colored in blue) and DFCI (colored in red) results are plotted above and below the zero line, respectively. Filled and open highlighted circles denote associations passing FDR < 0.05 and nominal P < 0.05, respectively. **c,** Proportion of DRB1 residue positions located within the ARD among positions with and without FDR-significant associations. Enrichment was assessed using a one-sided Fisher’s exact test. **d,** Structural localization of the informative DRB1 positions 9, 10, 11, 12, 13, 32, 33, 37, 71, and 77 (in blue) in the crystal structure of HLA-DRB1*15:01 in complex with human myelin basic protein peptide (PDB ID: 1BX2). The DRB1 β-chain is shown in light blue, the HLA-DRA α-chain in grey and the bound peptide in yellow; groove-facing DRB1 residues are shown as sticks. **e,** Adjusted hazard ratios for amino-acid residues at DRB1 positions 11, 13 and 71, estimated using multivariable cause-specific Cox models as described in the Methods. Points and horizontal bars indicate hazard ratios and 95% confidence intervals, respectively; residues with nominal P < 0.05 are shown in blue. **f,** Amino-acid combinations at positions 11, 13 and 71 in DRB1 alleles with at least one risk-associated residue. Risk-associated residues are shown in blue, and the number of risk residues carried by each allele is indicated on the right. **g,** Cumulative incidence of adrenal insufficiency according to DRB1*15 dosage among first lines of therapy containing CTLA-4 inhibitor. Numbers of patients at risk are shown below the plot. **h,** Cumulative incidence of adrenal insufficiency among ICI-containing lines of therapy classified as high risk by the random survival forest model, stratified by the presence or absence of DRB1*15. LoT, line of therapy; RSF, random survival forest.

This observation motivated us to hypothesize that variation within *HLA-DRB1* antigen-recognition domain (ARD) could potentially shape the peptide-binding repertoire of *HLA-DRB1*, ultimately influencing which antigens are presented to T cells and thereby modulating risk of ICI-associated adrenal insufficiency. To test this hypothesis and further clarify the role of *HLA-DRB1* in the pathology of adrenal insufficiency, we performed a positional omnibus scan across all residues in the ARD of the *HLA-DRB1* mature protein sequence to identify amino acids within *HLA-DRB1* that mediate adrenal insufficiency risk^57^. DRB1 allele types of each patient were imputed from MSK-IMPACT-derived SNP data using HIBAG (see **Methods**), with a cohort-specific model trained on a subset of the cohort with matched whole-exome sequencing data (N= 3,748 patients) and validated against a non-overlapping subset with clinical grade HLA typing results (N= 141 patients) (**Figure S5a**). This allowed us to impute the HLA-DRB1 allele for a subset of 24,895 patients with a prediction accuracy on the test set of 0.92 (**Figure S5b**). For each residue position within the ARD, we compared a base Cox proportional hazard model of time from start of line of therapy (LoT) to the start of adrenal-insufficiency-positive LLM batch with demographic and clinical covariates alone as predictor variables, to a full model that additionally includes amino acid dosages at that position. Association significance was determined by likelihood ratio test using the fitted models. Ten positions in the ARD of *HLA-DRB1* reached the statistical significance at FDR < 0.05, suggesting that they are informative for the time-to-adrenal insufficiency risk estimation (**Figure 5b, Supplementary Tables 54–56**).

To assess whether these associated positions are functionally positioned to influence peptide presentation, we tested for enrichment of the FDR-significant positions within the known HLA-DRB1 peptide-binding groove. Using the published groove-lining residue set^58^, we observed significant enrichment of the association signals within the peptide-binding groove -indicating that the risk-associated positions are disproportionately concentrated at sites that directly contact bound peptide (p= 0.013, **Figure 5c,d**). Among those, positions 11 (FDR = 4.9×10^-4^), 13 (FDR = 5.4×10^-5^), and 71 (FDR = 1.6×10^-4^) emerged as the top-ranking positions by effect size, and associations at these positions were replicated in an external cohort from the Dana-Farber Cancer Institute (DFCI), where all three positions were associated with severe adrenal insufficiency (p = 5.6×10⁻⁴, 1.1×10⁻³ and 1.0×10⁻³ for positions 11, 13 and 71 respectively) (**Figure 5b, Supplementary Table 57**). Remarkably, these same HLA-DRB1 positions have been previously implicated in both rheumatoid arthritis and multiple sclerosis risk^57,59,60^, underscoring that the ICI association converges on a well-established autoimmune antigen-presentation axis.

At each of positions 11, 13, and 71, we identified specific high-risk amino acid residues from the fitted full Cox proportional hazard model, corresponding to proline at position 11, arginine at position 13, and alanine at position 71 (**Figure 5e, Supplementary Tables 58–60**). Notably, DRB1*15:01, DRB1*15:02, and DRB1*15:03 each carry the P-R-A configuration at all three risk positions 11-13-71, while other tested alleles carried at most two of the three risk residues (**Figure 5f, Supplementary Table 61**), suggesting that the DRB1*15 allele type drives this association with ICI-associated adrenal insufficiency risk. This same allele otherwise predisposes patients in general to multiple sclerosis^59,60^, suggesting a common predisposition pattern between multiple sclerosis and ICI-associated adrenal insufficiency.

To assess the clinical impact of DRB1*15 dosage on adrenal insufficiency risk, we examined cumulative incidence of adrenal insufficiency among 665 patients receiving their first CTLA-4 inhibitor containing regimens (89.2% in combination with PD-1). Patients carrying one copy of a risk allele (dosage 1; n=160; **Figure 5g, Supplementary Tables 62–64**) showed significantly higher cumulative adrenal insufficiency incidence at 13.9% (95% CI 7.9%-21.7%) compared to non-carriers (dosage 0, ref; n=472; p=1.0×10-2) at 6.7% (95% CI 3.9%-10.4%), and homozygous carriers (dosage 2; n=33) demonstrated the highest incidence (p=5.6×10^-7^), with 35.0% (95% CI 14.2%-56.8%) of homozygous carriers receiving anti-CTLA4 developing adrenal insufficiency within 12 months of the start of treatment. Subtype-specific analyses further indicated that this association was driven by secondary, rather than primary, adrenal insufficiency (*i.e.* hypophysitis): when cases were restricted to primary adrenal insufficiency (**Methods**), cumulative incidence did not associate with DRB1*15 dosage (p=0.53, **Figure S5c;** secondary adrenal insufficiency p= 5.64×10^-6^**, Figure S5d**). This restriction to secondary adrenal insufficiency aligns the genetic association with the characteristic pituitary toxicity of checkpoint blockade, particularly CTLA-4 inhibition, and supports a specific treatment-mediated autoimmune phenotype rather than a general predisposition to adrenal dysfunction. The association was specific to ICI-containing LoTs, as DRB1*15 dosage was not associated with adrenal insufficiency in non-ICI LoTs (p=0.135, **Figure S5e;** PD1/PDL1 inhibitor LoTs p= 5.26×10^-5^ **Figure S5f**), with no monotonic dosage pattern observed. These data demonstrate that each additional copy of DRB1*15 incrementally and substantially increased the risk of developing adrenal insufficiency following immune checkpoint blockade. Critically, the at-risk genotype is common: 31.1% of genotyped patients carried at least one DRB1*15 allele (25.9% one copy, 5.2% two copies; **Figure S5g**). A germline marker that identifies a third of ICI-treated patients as being at two-to five-fold elevated risk of adrenal insufficiency is within reach of routine pre-treatment testing, since HLA typing is already standard in other clinical settings.

Finally, we examined whether including germline information could refine our RSF risk models of adrenal insufficiency. Within the RSF-defined high-risk tertile (**Figure 3c**), DRB1*15 carrier status further stratified patients by cumulative adrenal insufficiency incidence. Within 12 months of the start of ICI therapy, high-risk DRB1*15 carriers experienced 1.85 times (12.0%) the incidences of adrenal insufficiency (6.5%) as non-carriers within the high-risk patient group (p=0.003, **Figure 5h).** Clinical features and germline variation are therefore complementary rather than redundant; DRB1*15 status carries information about adrenal insufficiency risk that routinely collected clinical variables do not capture and incorporating germline information improves toxicity prediction.

## Discussion

Oncologists are asked to weigh efficacy against toxicity, but the two sides of that judgment rest on very different evidence. For efficacy there is a deep and growing toolkit — molecular predictive biomarkers, multigene assays, and risk models built from routine clinical and laboratory data — while for toxicity there is almost nothing equivalent, and risk is still approximated from adverse event rates in trial arms drawn from populations with strict eligibility criteria. The MSK-Tox resource directly addresses this key gap in knowledge by systematically characterizing adverse event incidence, modeling adverse event risk, and mining inherited predispositions to adverse events across cancer types and treatment modalities. Together these establish the empirical basis for a new precision safety paradigm in oncology, wherein risk assessment for treatment harm can be built on the same clinical and genomic foundations that underpin existing precision oncology.

Previous efforts to phenotype treatment toxicity from the medical record have been limited by scale, scope, or reliance on structured data, constraining broader discovery. Early machine learning approaches analyzed only several hundred manually reviewed cases and achieved modest accuracy for individual events^61^, while structured data approaches have shown limited sensitivity for systemic therapy toxicities^62^, consistent with known limitations of laboratory values, medications, and diagnosis codes used in isolation^6–8^. More recent language model approaches have remained restricted to inpatient settings or cohorts of several hundred patients^63–65^. Across all of these efforts, toxicity phenotypes have served as endpoints in themselves rather than as a substrate for prediction, and none has been linked to pretreatment risk modeling or to tumor and germline sequencing in the way that precision oncology routinely links tumor phenotypes to treatment benefit. MSK-Tox extends these efforts by multiple orders of magnitude, capturing severity, timing, and clinician attribution across 2.77 million notes from 55,406 patients spanning an unselected pan-cancer population and all major treatment modalities, and pairing these phenotypes with routine laboratory values, tumor sequencing, and germline genotypes to enable predictive modeling and biomarker discovery.

This scale and scope of data are the basis for our discovery that toxicity risk is organ-and therapeutic context-specific rather than a general treatment liability. For example, CTLA-4 blockade increased liver, colon, and adrenal toxicity but not pneumonitis, and individual cancer types exhibited distinct toxicity signatures, including pneumonitis in lung cancer and liver injury in colorectal cancer. We also observed that immune-mediated toxicity is a poor general proxy for treatment benefit: rather than a consistent survival advantage, pneumonitis and liver toxicity were associated with inferior survival, particularly when severe, with similar associations among patients never treated with immunotherapy. Together, these findings illustrate how population-scale phenotyping of toxicity can address questions of incidence, clinical consequence, and biological determinants of toxicity that were inaccessible in smaller cohorts.

The same principle applied to risk prediction. Models trained on a pooled, toxicity-agnostic endpoint discriminated poorly among ICI-treated patients, whereas models trained on individual adverse events performed consistently better and separated patients into risk tertiles with clinically meaningful differences in absolute one-year incidence, more than doubling between low-and high-risk groups for ICI-associated liver toxicity. These models used only information available before the decision to treat, and adding pre-treatment laboratory values and tumor genomic features did not materially improve accuracy, suggesting that routinely collected demographic, disease and treatment variables already capture most of the pretreatment signal accessible from the medical record. Feature attribution indicated why a single pooled model underperforms: the variables driving predicted risk differ between adverse events, and for sex and age they reverse in direction within a single cancer type.

Importantly, when considering all adverse events in aggregate (*i.e.* agnostic to the target organ), we found no germline variants that associated significantly with risk of developing an adverse event. In contrast, when considering both the target organ and the therapeutic context, associations with germline loci appeared. Some loci reflected vulnerability of the affected organ itself, including a variant near *FOXE1* implicated in thyroid development that is associated with risk of hypothyroidism regardless of whether a patient received immunotherapy. This implies that cancer treatment may unmask a pre-existing subclinical predisposition to thyroid dysfunction mediated by *FOXE1*. Other germline associations are conditional on the therapy: the strongest signal, at *HLA-DRB1*, appeared only in checkpoint-treated patients and resolved to specific residues lining the groove of *HLA-DRB1* that presents antigen. Interestingly, the same positions have been previously implicated in rheumatoid arthritis and multiple sclerosis^57,59,60^. These loci build on chemotherapy pharmacogenomics, where DPYD genotyping is now performed routinely before fluoropyrimidine treatment and is used to set the starting dose^66^. DPYD identifies a drug-specific risk of toxicity in general, whereas the associations described here depend on the specific toxicity and its treatment context, and reflect organ vulnerability or immune recognition rather than drug metabolism.

Our organ-agnostic composite analysis did not recover any of the three loci previously reported for pooled immune-related toxicities^43^. When the endpoint was resolved by organ and restricted to ICI-treated patients, however, two of the three showed nominal, directionally consistent associations — the IL7 locus with pneumonitis, and the 4p15 locus with hypothyroidism — neither of which was present in patients who did not receive ICI. One interpretation is that an association discovered under a pooled any-irAE definition is carried by a few constituent organ toxicities and diluted when these are aggregated with mechanistically unrelated events. Together, our findings argue that inherited susceptibility to cancer therapy toxicity is defined jointly by the organ at risk and the treatment given; a framing that explains why organ-agnostic searches have returned so little, and one that argues for organ and treatment-specific endpoints in future studies of treatment toxicity.

These findings have implications for trial design. For instance, in COSMIC-313, among patients with metastatic renal cell carcinoma, adding cabozantinib to nivolumab plus ipilimumab improved progression-free survival but raised grade 3–4 treatment-related toxicity from 43% to 75%^67^, and the final analysis showed no overall survival benefit^68^; restricting such intensified regimens to patients predicted to tolerate them could make escalation viable in populations where it currently is not. Conversely, de-escalation trials in the adjuvant setting could likewise weigh predicted long-term toxicity risk alongside ctDNA-defined recurrence risk^69^.

Taken together, these results argue that treatment harm has structure. Toxicity risk is not a diffuse property of intensive therapy but is organized by target organ and therapeutic context — an organization visible in incidence, prediction and germline architecture alike. Realizing precision safety in the clinic will require prospective and multi-institutional validation and extension beyond the six adverse events examined here, including for neurologic, cardiac, and nephrological toxicities. MSK-Tox provides the empirical foundation for that effort, and shows that the clinical and genomic infrastructure built to predict which patients will benefit from treatment can be turned, with equal rigor, to predicting which patients will be harmed by it.

## Methods

### Ethics approval

This study was approved by the Memorial Sloan Kettering Cancer Center Institutional Review Board protocol #26-175.

### Retrieval Augmented Generation system description

Clinical notes were pre-filtered using a retrieval-augmented generation (RAG) approach with FAISS indexing and BGE-large embeddings to retain only notes with semantic similarity to known toxicity descriptions. Row counts and total character counts were measured before and after the filtering step to quantify the reduction in data volume passed to downstream LLM classification.

The pipeline begins with a SageMaker Processing Job that ingests clinical notes (as CSV or Parquet), chunks them at sentence boundaries (∼200 tokens), and encodes all chunks using the BAAI/bge-large-en sentence transformer on GPU. The resulting embeddings are L2-normalized, indexed into a FAISS IndexFlatIP, and scored against predefined query terms for six irAE categories (adrenal insufficiency, colitis, hyperthyroidism, hypothyroidism, pneumonitis, liver toxicity). Chunks exceeding a cosine similarity threshold of 0.80 for any category are retained, producing a filtered CSV of relevant clinical text with per-category similarity scores. This job runs on G5 or G6 GPU instances (replacing the now-deprecated P3 fleet), with multi-GPU support for production-scale runs of 1000+ patients.

The filtered chunks are then processed and grouped by patient and 90-day temporal windows, concatenating chunk text with date headers into batches suitable for LLM input. Each batch represents one patient-window and carries metadata including the toxicity categories detected during retrieval and an estimated token count.

These batches are uploaded to S3 and processed through an AWS Step Functions state machine that orchestrates three Lambda functions: a splitter that partitions the CSV into parallel-processable chunks, a processor that calls Llama 4 Maverick 17B on Bedrock for each patient-window to output per-toxicity confidence scores (0–1) with rationale, and an aggregator that merges all results into a final CSV. The Map state runs up to 10 chunks concurrently with retry logic and fault isolation, so individual failures don’t block the pipeline.

### Toxicity annotation

Each patient-window batch was submitted to Llama 4 Maverick 17B with a single detection prompt returning a continuous confidence score between 0 and 1 for each of the six adverse events, together with a free-text rationale, constrained to a JSON object with six confidence fields and one rationale field. The prompt supplied a definition of each toxicity written to require attribution to systemic anti-cancer therapy, and paired each definition with explicit exclusions: infectious causes, pre-existing or intrinsic organ disease, direct tumor involvement of the affected organ, and toxicity arising from prior surgery or prolonged corticosteroid exposure were scored zero unless the note specified that systemic therapy had exacerbated them. A confidence score above zero required clear evidence that the toxicity was present at the time the note was written; findings mentioned only as a differential diagnosis or an anticipated risk, findings explicitly ruled out, and toxicities described as historical, resolved, improved or treated were scored zero. Nonspecific symptoms such as fatigue, and imaging findings readily explained by other causes, were specified as insufficient on their own; the model was instructed that all six toxicities could legitimately be absent from a note, and to score conservatively where evidence was ambiguous. The rationale field required the model to cite the specific evidence supporting or refuting each call and was retained for manual review. The full prompt is reproduced in **Supplementary Note 1**. Batches were called positive for a given toxicity when the returned confidence score exceeded the threshold determined by ROC analysis (see ROC curve analysis for optimal confidence determination).

Five of the six adverse events correspond to single CTCAE v5.0 terms (adrenal insufficiency, colitis, hyperthyroidism, hypothyroidism, pneumonitis). Liver toxicity has no equivalent single term and was defined as a composite of the CTCAE v5.0 laboratory terms for hepatocellular and cholestatic injury — alanine aminotransferase increased, aspartate aminotransferase increased, blood bilirubin increased, alkaline phosphatase increased and gamma-glutamyl transferase increased — which CTCAE grades separately but which are reported here as one endpoint, since drug-induced liver injury is not captured by any one of them in isolation.

### Grade and subtype annotation

Toxicity detection and toxicity characterization were performed as separate passes with separate prompts. Batches returning a positive call for a given toxicity were re-submitted to a second, toxicity-specific prompt that presented the corresponding CTCAE grading criteria alongside the retrieved clinical text for that patient-window and returned an imputed CTCAE grade (**Supplementary Note 2**); batches with no positive call for that toxicity were not graded. For liver toxicity, the grading prompt presented the criteria for each constituent CTCAE v5.0 term and the batch was assigned the highest grade reached by any of them.

For adrenal insufficiency, a further prompt was applied to positive batches to distinguish primary adrenal insufficiency, arising from direct adrenal injury, from secondary (central) adrenal insufficiency arising from hypophysitis (**Supplementary Note 3**). This distinction cannot be made from the detection call alone and is mechanistically relevant in the immune checkpoint inhibitor setting, where hypophysitis is a recognized cause of central adrenal insufficiency.

#### Gold standard dataset description

We compared the performance of Tox-Annotator for the 6 adverse events to the gold standard clinical trial-based dataset. This was derived from a prospectively collected gold standard dataset of structured Common Terminology Criteria for Adverse Events (CTCAE), consisting of 6,279 patients from across 1,057 individual clinical trials between 1/1/2010 and 7/29/2024 at MSK. The dataset includes adverse event type, date of occurrence, grade, and adverse event attribution.

### ROC curve analysis for optimal confidence determination

We used a confidence-based approach to optimize Tox-Annotator AE calls. Specifically, we designed a prompt to output the confidence of the LLM in calling each of the 6 AEs of interest. We then used a subset of 1000 patients from the gold standard dataset, divided into 700 patients of training and 300 patients for testing, to determine the optimal operating threshold for each toxicity to maximize the F1 score on the training set and evaluating them on the test set. We manually reviewed all discrepancies between Tox-Annotator and the CTCAE-based gold standard to adjudicate the source of error across all 1000 patients. This confidence-based approach improved performance compared to the binary approach for each of the 6 AEs (**Figure S1f**). The optimal operating threshold for each toxicity was selected to maximize the F1 score on the training set and evaluating them on the test set, leading to different thresholds for each toxicity. AUC was computed using the trapezoidal rule. Test-set metrics are reported with 95% confidence intervals from 2,000 stratified bootstrap resamples, as above.

As previously described^70^, each discrepant assessment was manually reviewed using the corresponding clinical documentation to determine whether the discrepancy reflected an error in the Tox-Annotator classification or in the CTCAE-based gold-standard annotation. Of the 197 discrepancies found in the held-out test set, 87 (44.2%) were adjudicated as errors in the CTCAE-based annotation rather than the Tox-Annotator classification. Performance metrics were subsequently recalculated using the manually adjudicated classifications. AUC values in the held-out test set were 0.97 (95% CI, 0.95–0.98) for pneumonitis, 1.00 (95% CI, 1.00–1.00) for adrenal insufficiency, 0.81 (95% CI, 0.76–0.87) for liver toxicity, 0.92 (95% CI, 0.83–0.99) for colitis, 0.77 (95% CI, 0.56–0.98) for hyperthyroidism, and 0.90 (95% CI, 0.86–0.95) for hypothyroidism. F1 scores ranged from 0.40 to 0.90 across adverse events (**Figure 1b**, **Figure S1e, Supplementary Tables 4 and 65**). False negatives and false positives in **Figure 1b** are partitioned by adjudicated source, pipeline error versus gold-standard curation error, determined by comparing the original and post-adjudication gold standards. Adjudication of the source of error was done by three reviewers (Z.B, S.M., A.O.) and disagreements were resolved using the consensus method.

### Independent validation set

Gold-standard patients not included in the 1,000-patient train/test subset (n = 5,279) were scored using the institution-wide pipeline run, applying the toxicity-specific thresholds locked from the training set without further modification. No manual adjudication of discrepancies was performed in this set, so performance is reported against the original, uncorrected gold standard. Confidence intervals were obtained from 2,000 bootstrap resamples.

### Pre versus Post-RAG comparison

The LLM classification pipeline was run on both the full (pre-RAG) and RAG-filtered (post-RAG) clinical note datasets for each of the six AEs. F1 scores were computed for each condition, with 95% bootstrap confidence intervals obtained by drawing 2,000 stratified bootstrap resamples, computing F1 at the optimal threshold for each resample, and taking the 2.5th and 97.5th percentiles. The same model and prompt were used for both cases. The optimal operating threshold for each toxicity was selected to maximize the F1 score for each case separately. Pre-and post-RAG costs were estimated from an observed post-RAG LLM inference cost, scaled by the ratio of input-text character counts per patient before versus after retrieval filtering. Post-RAG cost additionally included the compute cost of embedding and indexing (the RAG run). Both the LLM and SageMaker components were extrapolated to 55,406 patients; pre-RAG cost comprised LLM inference only.

### Confidence-based versus binary comparison

F1 scores were computed for each AE under two LLM output configurations: a confidence-based mode, in which the model returned a continuous confidence score that was thresholded at the optimal operating point from the ROC analysis, and a binary mode, in which the model was prompted to return a direct yes/no classification. Error bars represent 95% bootstrap confidence intervals, constructed by drawing 2,000 stratified bootstrap resamples, computing F1 at the optimal threshold for each resample, and taking the 2.5th and 97.5th percentiles of the resulting distribution. All comparisons used the same underlying test dataset, RAG queries, and prompt structure.

### Comparison across different LLM models

Four models (Llama 4 Maverick, Llama 4 Scout, Claude Sonnet 4.5, GPT-OSS-120B) were run with identical RAG retrieval, prompts and inputs on a 300-patient subset split 149/151 into threshold-selection and evaluation sets. Toxicity-specific thresholds were optimized for F1 in the 149-patient set and applied unchanged to the evaluation set. Confidence intervals used 2,000 stratified bootstrap resamples, and each model was compared with Llama 4 Maverick by two-sided paired bootstrap test. Per-model cohort inference cost was estimated by scaling each model’s observed per-token pricing to the full cohort to a 70,000-patient reference run.

### LLM grade validation

LLM-imputed grade was compared to gold standard grade by subtracting gold-standard CTCAE grade from LLM-imputed grade for each matched patient–toxicity comparison. Exact-grade agreement was defined as the proportion of comparisons in which the LLM-imputed and gold-standard grades were identical. Statistical significance of exact-grade agreement was assessed using a permutation-based null model in which LLM-imputed grades were randomly shuffled across matched comparisons while gold-standard grades were held fixed. The null distribution was generated using 5,000 permutations, and the one-sided *P* value was calculated as the proportion of permuted exact-match rates greater than or equal to the observed exact-match rate.

### LLM time-to-event validation

Time-dependent predictive performance of LLM-derived adverse event probabilities was evaluated using the time-integrated Brier score over a fixed 0–12-month period from the start of each line of therapy. For each toxicity, LLM probabilities were matched to the corresponding patient and line of therapy, and gold standard event times were defined relative to the start of that line, with follow-up censored at the line-specific censoring time. The time-integrated Brier score was calculated from binary event-by-time status after applying the toxicity-specific thresholds. Ninety-five percent confidence intervals were estimated using 500 bootstrap resamples of lines of therapy. Statistical significance was assessed using a permutation-based null model generated by randomly shuffling LLM probabilities across lines of therapy while holding gold-standard outcomes fixed. One-sided P values were calculated as the proportion of 10,000 permutations with Brier scores less than or equal to the observed Brier score.

### Laboratory validation of toxicity calls

For each patient and toxicity, a reference date was defined as the start of the first 90-day window attaining that patient’s maximum LLM-imputed grade; for patients without the toxicity (grade 0), the midpoint of their annotated timeline was used. Within each window, a given analyte was summarized as a time-averaged area under the concentration–time curve (trapezoidal integration over observed results, divided by window length in days) for 60-day windows starting with the start dates described above; patients with a single result in the window contributed that value. Analytes were mapped to toxicities a priori (cortisol, adrenal insufficiency; TSH, hypothyroidism; ALT, liver toxicity) after harmonizing laboratory test names. Grades with fewer than 5 patients were merged with the adjacent grade. Monotonic trend across grades was tested with the Jonckheere–Terpstra test, and adjacent grades were compared with two-sided Mann–Whitney U tests.

### Steroid exposure validation of toxicity calls

Reference dates were defined as above. Pre-and post-toxicity windows were defined as the 60 days before and after the reference date. Within each window, cumulative corticosteroid exposure was computed in prednisone-equivalent mg-days using standard equivalence factors (hydrocortisone 0.25, cortisone 0.20, prednisone and prednisolone 1.0, methylprednisolone and triamcinolone 1.25, fludrocortisone 2.5, dexamethasone 6.67, betamethasone 8.33). Analyses were restricted to lines of therapy containing an immune checkpoint inhibitor and to patients with any corticosteroid exposure in either window. The outcome was log_2_((post + 0.01)/(pre + 0.01)). Monotonic trend across grades was tested with the Jonckheere–Terpstra test, and adjacent grades were compared with two-sided Mann–Whitney U tests.

### Censoring and follow-up

Time zero was defined as the start of the index line of therapy. Unless otherwise indicated, patients were censored at the earliest of the day before initiation of a subsequent line of therapy, death, 180 days after completion of the index line of therapy, or the last documented clinical follow-up. This censoring strategy was designed to attribute adverse events to the index treatment exposure while minimizing attribution to subsequent therapies. Patients who did not experience the adverse event of interest by the censoring date were censored. Median follow-up was estimated using the inverse Kaplan-Meier method.

### Treatment regimen and modality assignment

Medication administration records were first cleaned by imputing missing end dates from the corresponding start date, correcting records whose end date preceded the start date, and extending zero-duration administrations by one day; oral and topical agents were processed as a separate stream from parenteral agents. For each patient and each generic agent, consecutive administrations were collapsed into continuous exposure intervals, with a new interval initiated whenever the interval between the end of one administration and the start of the next exceeded 90 days. Drug-level intervals were then resolved into regimens: for each patient, all agent intervals were placed in an interval tree and split at every interval boundary, so that each resulting time segment was defined by the specific set of agents administered concurrently during that segment. Each regimen was annotated according to the treatment classes it contained — chemotherapy, immunotherapy, targeted therapy, biologic, and hormonal therapy — based on agent-level class assignments. Lines of treatment were derived from records of administered treatment regimens, each capturing one or more drugs given together over a defined start and end date. Because the same ongoing regimen was often split across multiple entries, adjacent entries separated by 30 days or less and sharing at least one drug were merged into a single continuous treatment cycle. Regimens not classified as chemotherapy, immunotherapy, biologic, targeted, hormonal, or investigational therapy were then excluded, and the remaining episodes were ordered chronologically within each patient to assign a sequential line of treatment.

### Comparisons of toxicity incidence across treatment, cancer type, sex and age

Differences in toxicity incidence by systemic therapy class, cancer type, sex, and age group were assessed using a common approach. For each comparison, one-year cumulative incidence was estimated separately within each group, and groups were then compared using multivariable Cox proportional-hazards models fitted independently for each toxicity, with a light ridge penalty applied as a safeguard against near-separation in sparse strata. The exposure of interest was entered as a binary indicator against a fixed reference group: non-ICI regimens for the ICI comparison; PD-(L)1 without CTLA-4 for the within-ICI comparison; all other cancer types for each cancer type, in a one-versus-rest design; male sex; and age under 50 years. Models were adjusted for systemic therapy composition (PD-1/PD-L1 and CTLA-4 exposure, and concomitant chemotherapy, hormone, biologic and targeted therapy), age, sex and cancer type, except that any covariate defining the exposure in a given comparison was omitted from that model. Binary covariates with a zero-event cell were removed, and remaining adjustment covariates were dropped in a pre-specified order until at least ten events per estimated parameter remained; the exposure indicator was never dropped. Models were fitted on complete cases with respect to age, sex, cancer type and follow-up time, with missing systemic therapy flags treated as absent; patients with more than one primary cancer type were additionally excluded from the cancer type analysis. Reported p-values are Wald tests on the exposure coefficient.

### Line-of-therapy analyses

The unit of analysis was the line of therapy rather than the patient, with lines grouped as 1, 2, 3 and 4 or later. One-year cumulative incidence was estimated within each group as above. Because patients contribute multiple lines, inferential comparisons used a shared-frailty Cox proportional-hazards model fitted in R with coxme, specifying a patient-level random intercept on the log-hazard scale. Fixed effects were line-of-therapy group (reference, line 1), the PD-(L)1 and CTLA-4 agent flags, concomitant chemotherapy, hormone therapy, biologic therapy and targeted therapy, mean-centered age at line start, sex (reference, male) and cancer type (reference, “Other”). Hazard ratios and 95% confidence intervals were derived from the fixed-effect estimates and their model-based standard errors, and the estimated frailty variance is reported for each toxicity.

### Association of toxicity with progression-free and overall survival

Time zero was the start date of the first line of therapy. Progression-free survival was defined as the time to the earlier of first radiologically determined progression or death, and overall survival as the time to death; both were censored at last documented clinical follow-up. Radiological progression was taken from an NLP-derived timeline of imaging reports, with reports labelled as progression or indeterminate for progression counted as events. Each toxicity was modelled as a time-varying exposure: a patient’s follow-up was split at the toxicity onset date into an unexposed interval before onset and an exposed interval from onset onward, with patients whose onset fell at or after their event or censoring time contributing a single unexposed interval. Models were fitted with lifelines (CoxTimeVaryingFitter, penalizer = 0.1), stratified by cancer type so that each type carries its own baseline hazard, and adjusted for age, sex, body mass index and concomitant chemotherapy, targeted therapy, hormone therapy and biologic therapy on the index line. Cancer type strata containing fewer than two patients were excluded. Analyses were run separately for grade 1 or higher and grade 3 or higher toxicity; grade 1+ onset was taken as the first window exceeding the toxicity-specific confidence threshold, and grade 3+ onset as the first window with an imputed CTCAE grade of 3 or higher. Models were fitted only where at least ten outcome events and both exposure states were observed, and results are displayed only for toxicities with at least ten exposed patients. All three treatment cohorts, patients whose first line contained an ICI, patients whose first line did not, and all patients combined, were fitted; the figures display the ICI cohort, and the remaining cohorts are reported in the supplementary tables.

### Toxicity grade trajectories around severe events

Patients reaching grade 3 or higher for a given toxicity were identified, and the distribution of that toxicity’s grade was tabulated at successive 90-day windows relative to the severe episode. Pre-event offsets (−180 and −90 days) were anchored to the patient’s first grade 3+ window and post-event offsets (+90 and +180 days) to their last grade 3+ window, with the peak episode placed at day 0. To hold the denominator constant across offsets, only patients with observed window data at every displayed offset were included. Hyperthyroidism was not analyzed because too few grade 3+ events were observed for a stable trajectory.

### Random Survival Forest Model

Patients features consist of age, self-reported sex, cancer staging (stage 1-3 vs. stage 4), histopathology-based cancer type, and sequencing-derived genetic ancestry. Smoking status was derived from clinical notes using natural language processing models, as previous described^70^. Lab values from CBC and CMP obtained in the 7 days before treatment start was used to calculate min/max features. The analyte panel comprised: ALT, AST, anion gap, basophils, calcium, carbon dioxide, chloride, creatinine, eosinophils, glucose, hematocrit, hemoglobin, immature granulocytes, large unstained cells (LUC), lymphocytes, MCH, MCHC, MCV, monocytes, neutrophils, nucleated erythrocytes, platelets, potassium, RBC, RDW, sodium, urea nitrogen, and WBC. Tumor genomic features consist of tumor mutation burden, fraction genome altered, and microsatellite instability score derived from MSK-IMPACT sequencing.

Random survival forests (RSF)^71^ models were trained to predict time from treatment initiation to adverse event onset. Models were trained independently on each line of therapy given to a patient, right censored at the start of a subsequent line of therapy, time of death, or time of last follow-up. Adverse event–specific models were trained separately for each of the six adverse events using grade 1+ incidence as the endpoint. In addition, toxicity-agnostic models were trained on the onset of any of the six pre-specified adverse events, at both grade 1+ and grade 3+ thresholds. Models were trained using the scikit-survival (https://github.com/sebp/scikit-survival) package. All models use the same pre-determined hyperparameter configuration (number of trees = 3000; square root rule for feature subsampling per split; minimum samples per leaf = 25; minimum samples to split an internal node = 50; maximum tree depth unconstrained; and bootstrap sampling enabled). Train-test split was created at the patient level with 80/20 train/test ratio. Model performance was measured using Harrell’s concordance index (c-index). Post-hoc feature attribution was performed using SHAP^72^.

### Genome-wide association analysis

We performed time-to-event genome-wide association studies to identify germline variants associated with systemic therapy-associated adverse event risk. Analyses used hard-called imputed germline genotypes. Patients were analyzed in treatment-context cohorts defined by line of therapy: the combined cohort was anchored at the first observed line of therapy, the ICI cohort was anchored at the first line of therapy containing immunotherapy, the non-ICI cohort was anchored at the first line of therapy containing systemic therapies other than immunotherapy. For each index line, time zero was defined as the line-of-therapy start date. Patients were censored at the earliest of line-of-therapy end plus 180 days, the day before the next line of therapy, death, or last follow-up.

### Adverse event phenotypes

Genome-wide analyses focused on individual all-grade adverse event phenotypes derived from MSK-Tox annotations: pneumonitis, adrenal insufficiency, hypothyroidism, hyperthyroidism, colitis, and liver toxicity. Adverse event onset was defined as the first LLM-positive temporal window beginning on or after time zero and before censoring. Patients without an adverse event before censoring were treated as censored observations.

### Germline statistical model

GWAS was performed using Cox proportional hazards score tests implemented in SPACox. For each adverse event, survival time was calculated from the index line of therapy start to adverse event onset or censoring. Models were adjusted for age at line of therapy start, sex, line of therapy number, treatment composition covariates, cancer type indicator variables, the first ten genetic principal components, and MSK-IMPACT panel version. Treatment covariates included indicators for chemotherapy, CTLA-4 therapy, PD-1 therapy, biologic therapy, and targeted therapy. Cancer type was modeled using indicator variables for the most frequent cancer types. Genotype samples were ordered to match the PLINK family file before fitting the SPACox null model and running genome-wide tests. Genome-wide significance was defined as P < 5 × 10^-8, and suggestive associations were summarized at P < 1 × 10^-5. Genomic inflation was assessed using lambda genomic control inflation factor.

### Replication of the germline findings in the DFCI cohort

We sought to replicate genome-wide significant associations in an independent cohort of 6,112 patients (4,989 for EUR) ICI-treated patients from the Dana-Farber Cancer Institute (DFCI) PROFILE cohort with imputed germline genotypes. For each adverse event, a parallel time-to-event genome-wide association analysis was performed using the same SPACox Cox proportional-hazards score-test framework as the discovery analysis, with analogous covariate adjustment for age, sex, PD-1 treatment (1/0), PD-L1 treatment (1/0), CTLA-4 treatment (1/0), receipt of other therapies (e.g., chemotherapy; 1/0), cancer type, the first 10 genetic principal components, tumor sequencing panel version, and whether tumor sequencing was performed after treatment initiation (1/0). In addition to the specific adverse events, broader ICD-and CTCAE-defined endocrine and respiratory disease categories were analyzed, each at graded severity thresholds.

Because an individual lead variant may be absent from, or under-powered in, the replication cohort, replication was assessed using a linkage-disequilibrium (LD) proxy approach. For each MSK-Tox lead variant, proxy variants were defined as SNPs within ±500 kb and in LD at r² ≥ 0.6 with the lead variant, identified with PLINK2 (--r2-unphased, --ld-snp-list, --ld-window-kb 500, --ld-window-r2 0.6). LD was computed in two reference panels and the resulting proxy sets were merged: (i) the DFCI PROFILE cohort itself (in-sample LD, restricted to the analyzed European-ancestry samples or all individuals), and (ii) 1000 Genomes Phase 3 European-ancestry reference samples or EUR + AFR-ancestry reference samples; candidate proxies were de-duplicated by genomic position and annotated by source (PROFILE, 1000 Genomes, or both).

Lead variants and proxies were defined on GRCh37/hg19 and matched to PROFILE and 1000 Genomes variant identifiers by chromosome and position.

For each lead locus, all candidate proxies present in the DFCI association results for the corresponding phenotype were examined, and the DFCI association statistics (hazard ratio and P value) were retrieved for the proxy with the strongest DFCI association (”best proxy”); the LD (r²) between this proxy and the lead variant is reported alongside its DFCI P value. Replication was considered nominal at P < 0.05 and genome-wide significant at P < 5 × 10⁻⁸ for the matched phenotype.

### External validation in population biobanks

For each genome-wide significant lead variant, we additionally queried two external population-scale biobanks—FinnGen and UK Biobank—to assess whether the locus is associated with the corresponding organ-specific disease phenotype in the general population. Because these are general-population resources that do not capture treatment-emergent immune-related or systemic-therapy toxicities, look-ups were directed at the cognate disease endpoints for each locus (thyroid disease for the *FOXE1* [rs10983761]; interstitial-lung and respiratory disease for the *FOXP2* [rs12705907] locus; adrenal and autoimmune endpoints for the *HLA-DRB1* [rs112463084] locus). Lead variants were defined on GRCh37/hg19 and converted to GRCh38 coordinates using the Ensembl REST API prior to look-up, as both resources are built on GRCh38; effect direction was recorded relative to the discovery risk allele.

### FinnGen

Each lead variant was queried against FinnGen Data Freeze 12 (R12) using the public per-variant phenome-wide association interface, retrieving association statistics across registry-defined disease endpoints. The *HLA-DRB1* lead variant (rs112463084) was not evaluable in FinnGen because it failed FinnGen variant quality control (low imputation call rate); the HLA region was instead resolved by allele-level imputation and positional testing for further validation (see below). The remaining lead variants (rs10983761, rs112386546, rs12705907) were present and evaluated.

### UK Biobank

UK Biobank GWAS summary statistics are not available through a public per-variant query interface, we therefore obtained UK Biobank associations through the Open Targets Platform GraphQL API, which harmonizes published GWAS—including UK Biobank–based studies—into fine-mapped credible sets annotated by contributing cohort, retaining associations from UK Biobank–derived studies. Because the Open Targets variant index contains only variants residing within a fine-mapped credible set, only the common *FOXE1* lead variant (rs10983761) was indexed and evaluable; the other lead variants (rs112386546, rs12705907, rs112463084) were not present in the index and could not be evaluated by this approach.

### Fine–Gray subdistribution hazard models

For each genome-wide significant lead variant, we estimated its association with the corresponding adverse event separately within ICI-treated patients, non–ICI-treated patients, and the combined cohort. In each group we used the same event definition and censoring as the genome-wide analysis (time from first line-of-therapy start; censoring at the earliest of line end + 180 days, next line start - 1 day, last follow-up, or death) but treated death as a competing event, and fit Fine–Gray subdistribution hazard models with the lead variant modeled as a continuous imputed allele dosage (expected effect-allele count, 0–2), adjusting for age at line-of-therapy start, sex, and line of therapy number (where appropriate). Per-allele subdistribution hazard ratios with 95% confidence intervals are reported. Hazard ratios are reported per copy of the alternate allele on GRCh37.

### HLA-DRB1 Genotype Imputation

HLA-DRB1 allele imputation was performed using HIBAG (HLA Imputation for Biomedical Attribute Genetics) with GPU acceleration via the *HIBAG.gpu* package. A locus-specific classifier ensemble was trained on samples with available whole-exome sequencing-derived HLA genotypes determined by HLA-HD, using PLINK-format genotype data as input. SNPs within a 500 kb flanking window of DRB1 were selected using *hlaFlankingSNP*, and alleles were standardized to two-field resolution. Two independent classifier ensembles of 100 classifiers each were trained on GPU and subsequently merged using *hlaCombineModelObj,* yielding a final ensemble of 200 classifiers. Model accuracy was evaluated in an independent validation cohort with HLA genotypes determined by Histogenetics clinical typing. Imputed calls were compared to ground-truth alleles across a range of HIBAG posterior probability thresholds (0.00-0.95), using amino acid group concordance as the accuracy metric. Concordance was defined based on identical antigen recognition domain amino acid sequences derived from IPD-IMGT/HLA, such that alleles sharing the same DRB1 peptide-binding groove sequence were considered equivalent regardless of synonymous or intronic variation. A posterior probability threshold of 0.35 was selected for HLA-DRB1, at which two-field imputation accuracy reached 92%. Imputation was then applied to the full pancancer GWAS cohort, retaining only calls meeting this confidence threshold for downstream analyses.

### Positional testing of HLA-DRB1 mature protein sequence for adrenal insufficiency associations

To identify informative amino acid positions within HLA-DRB1, we performed a position-wise omnibus association scan using Cox proportional hazards models. For each DRB1 mature protein’s antigen recognition domain position, inferred allele pairs were converted into amino acid dosage variables, representing the number of copies of each residue carried by a patient. Monomorphic positions were excluded. At each polymorphic position, the most common residue was used as the reference category and removed from the model to avoid collinearity. For each grade threshold, we analyzed patients receiving a first line of therapy containing immunotherapy. Patients with multiple cancer diagnoses, unknown sex, non-mapped ancestry, age <18 years at treatment initiation, or missing covariates were excluded. The base Cox model adjusted for age at treatment start, sex, ancestry, treatment covariates, and cancer type, with cancer type included as a stratification variable in pan-cancer analyses. In the full model, residue dosage terms for a given DRB1 position were then added jointly to the base model. Evidence for association at each position was evaluated using a likelihood ratio test comparing the full model containing residue dosage terms to the base model. Robust variance estimation was used for Cox model fitting. P values from the position-wise omnibus tests were corrected across tested DRB1 positions using the Benjamini-Hochberg false discovery rate procedure. Positions passing FDR < 0.05 were considered informative and were further inspected using residue-level effect estimates and forest plots.

### DFCI validation of HLA findings

HLA alleles at seven classical loci were independently imputed in the DFCI cohort from chromosome 6 genotype data using SNP2HLA v1.0.3. Variants with an imputation INFO score >0.4 were retained, converted to PLINK binary format, and harmonized to 1000 Genomes Project Phase 3 variant identifiers. Samples were divided into approximately 100 batches for parallel imputation. For HLA-DRB1, four-digit allele dosages were rounded to derive two inferred alleles per individual, and samples without exactly two called alleles were excluded. Among 4,989 individuals in the original European-ancestry phenotype cohort, 3,893 had available HLA-DRB1 calls. For each phenotype, the Cox model sample comprised the overlap between the corresponding ancestry-and phenotype-specific cohort and individuals with an available HLA-DRB1 call and complete covariate data.

To validate the HLA-DRB1 positional associations identified in the primary cohort, imputed HLA-DRB1 alleles were mapped to amino-acid sequences in the antigen-recognition domain using IMGT mature-protein coordinates, and residue dosages (0, 1, or 2 copies) were calculated at each polymorphic position. Associations between each amino-acid position and time to immune-related adverse event onset were evaluated using likelihood-ratio tests comparing Cox proportional-hazards models with and without residue-dosage terms for that position. Follow-up began at initiation of first-line immune-checkpoint inhibitor therapy, and patients without an event were censored at the earliest of death, loss to follow-up, or initiation of a second line of immune-checkpoint inhibitor therapy. Models were adjusted for cancer type, age, sex, treatment start year, tumor-sequencing panel version, whether tumor profiling occurred after treatment initiation, receipt of PD-1, PD-L1, or CTLA-4 therapy, concurrent treatment, and the first 10 genetic principal components. Analyses were performed separately in the genetically inferred European-ancestry cohort and in the overall multi-ancestry cohort.

### Structural visualization of HLA-DRB1*15 protein

To illustrate the structural context of the DRB1 risk-associated amino acid positions, the crystal structure of HLA-DRB1*15:01 in complex with human myelin basic protein peptide (PDB ID: 1BX2) was visualized using UCSF *ChimeraX*. The peptide-binding groove was rendered in surface representation in grey, and the bound peptide was displayed in cartoon representation in yellow. Residues at positions 9, 10, 11, 12, 13, 32, 33, 37, 71, and 77 of the mature DRB1 protein -- which are shared across the risk-associated alleles DRB1*15:01, *15:02, and *15:03 -- were displayed as sticks and colored in blue to highlight their location within the groove. Numbering follows the IPD-IMGT/HLA mature protein convention. The 1BX2 structure was selected as a representative model for the DRB1*15 risk haplotype; the co-crystallized myelin basic protein peptide is not implicated in ICI-associated adrenal insufficiency and is shown solely to illustrate peptide-binding groove architecture.

## Data and code availability

All data underlying the figures and reported statistics are provided in Supplementary Tables 1–65. All code necessary to reproduce the findings in the manuscript are shared at https://github.com/ziadbakouny18/msk-tox. Raw sequencing data are restricted to protect patient privacy in accordance with applicable federal and state laws and institutional policies. Additional de-identified individual-level clinical data may be made available from the corresponding authors upon reasonable request and subject to institutional review and execution of an appropriate data use agreement.

**Supplementary Table 1.** F1 scores before and after retrieval-augmented filtering in 110 patients. Per-toxicity pre-and post-RAG F1 with 95% confidence intervals from 2,000 bootstrap resamples, the paired difference, and two-sided paired bootstrap P values. Corresponds to **Figure S1c**.

**Supplementary Table 2.** Estimated inference cost before and after retrieval-augmented filtering, extrapolated to 55,406 patients. Input-text characters per patient pre-and post-RAG and their ratio, LLM inference cost under each condition, embedding and indexing compute cost, total post-RAG cost, and absolute and percentage savings. Corresponds to **Figure S1d**.

**Supplementary Table 3.** Estimated inference cost for a 70,000-patient run by model for Llama 4 Maverick, Llama 4 Scout, Claude Sonnet 4.5 and GPT-OSS-120B, with cost expressed relative to the least expensive model. Corresponds to **Figure S1h**.

**Supplementary Table 4.** Performance of Tox-Annotator against the CTCAE-based gold standard in the 300-patient held-out test set. Sensitivity, specificity, precision, F1 and AUC for each of the six adverse events, at the toxicity-specific operating threshold selected in the training set, with 95% confidence intervals from 2,000 bootstrap resamples. Corresponds to **Figure 1b**.

**Supplementary Table 5.** Performance of Tox-Annotator in the independent validation set of 5,279 gold-standard patients outside the 1,000-patient train/test subset. Counts (TP, FP, TN, FN), sensitivity, specificity, precision, F1 and AUC with 95% bootstrap confidence intervals, evaluated against the original gold standard without adjudication of discrepancies.

**Supplementary Table 6.** Agreement between Tox-Annotator-imputed and gold-standard adverse event grade across 1,454 matched patient–toxicity comparisons. Count and percentage of comparisons at each signed grade difference (LLM minus gold standard), the observed exact-match rate, the mean exact-match rate under a 5,000-permutation label-shuffled null, and the one-sided permutation P value. Corresponds to **Figure S1i**.

**Supplementary Table 7.** Time-dependent validation of Tox-Annotator adverse event probabilities by time-integrated Brier score over 12 months from the start of each line of therapy. Per-toxicity Brier score with 95% confidence intervals from 500 bootstrap resamples of lines of therapy, numbers of lines, patients and events, the prevalence-matched null Brier score with absolute and relative improvement, and the one-sided P value from a 10,000-permutation null. Corresponds to **Figure S1j**.

**Supplementary Tables 8–10**. Laboratory values by Tox-Annotator-imputed toxicity grade: cortisol in adrenal insufficiency (8, **Figure 1c**), TSH in hypothyroidism (9, **Figure 1d**), and ALT in liver toxicity (10, **Figure 1e**). Per-grade sample size, median and interquartile range of the time-averaged 60-day post-onset area under the curve on both the log2 and raw scales, adjacent-grade two-sided Mann–Whitney P values, and the Jonckheere–Terpstra trend statistic and P value across grades. Grades with fewer than five patients were merged with the adjacent grade.

**Supplementary Tables 11–13**. Corticosteroid exposure by Tox-Annotator-imputed toxicity grade for adrenal insufficiency (11, **Figure 1f**), colitis (12, **Figure 1g**) and pneumonitis (13, **Figure 1h**), among immune checkpoint inhibitor–treated patients with any steroid exposure. Per-grade sample size, median and interquartile range of log2(post/pre) prednisone-equivalent exposure in the 60 days before and after imputed onset, median post-onset exposure in mg-days, adjacent-grade Mann–Whitney P values, and the Jonckheere–Terpstra trend statistic and P value.

**Supplementary Table 14.** Distribution of first-line systemic therapy classes across the ten most common cancer types, corresponding to **Figure S2a**. Patient counts for each mutually exclusive treatment class, with row totals.

**Supplementary Table 15.** Distribution of CTCAE grades for toxicities occurring during first-line therapy, corresponding to **Figure S2b**. Number of patients at each grade for each toxicity.

**Supplementary Table 16.** Cohort size and event counts underlying the cumulative incidence curves in **Figure 2a**. Number of first-line patients at risk and number of first events for each of the six toxicities. Counts differ slightly between toxicities because patients with zero follow-up time for a given toxicity are excluded.

**Supplementary Table 17.** Numbers at risk for the cumulative incidence curves in **Figure 2a**. Patients remaining at risk for each toxicity at three-month intervals from the start of first-line therapy through 24 months.

**Supplementary Table 18.** One-year cumulative incidence of systemic therapy toxicities during first-line therapy, corresponding to **Figure 2b**. Kaplan–Meier estimates with 95% confidence intervals, ordered by descending incidence.

**Supplementary Table 19.** One-year cumulative incidence of systemic therapy toxicities during first-line therapy by sex, corresponding to **Figure S2c**. Kaplan–Meier estimates with 95% confidence intervals for each sex, with adjusted hazard ratios for female versus male from multivariable Cox models, the corresponding Wald p-values, unadjusted log-rank p-values, and Benjamini–Hochberg and Bonferroni-corrected p-values across the six toxicities.

**Supplementary Table 20.** One-year cumulative incidence of systemic therapy toxicities during first-line therapy by age group, corresponding to **Figure S2d**. Estimates with 95% confidence intervals for each age group, with adjusted hazard ratios relative to patients aged under 50 years, Wald and unadjusted log-rank p-values, and Benjamini–Hochberg and Bonferroni-corrected p-values across all toxicity-by-age-group comparisons. Columns documenting the estimator, covariate adjustment and multiplicity correction are included.

**Supplementary Table 21.** One-year cumulative incidence of systemic therapy toxicities by immune checkpoint inhibitor exposure, corresponding to **Figure 2c**. Estimates with 95% confidence intervals for ICI-treated and non-ICI-treated patients, with the adjusted p-value from the multivariable Cox model for each toxicity.

**Supplementary Table 22.** Full multivariable Cox proportional-hazards models for the ICI versus non-ICI comparison in **Figure 2c**. Hazard ratios, 95% confidence intervals and p-values for every term in each model, with cohort size, exposed count, event count, and any covariates removed by the zero-event-cell or events-per-parameter rules.

**Supplementary Table 23.** One-year cumulative incidence of systemic therapy toxicities by immune checkpoint inhibitor class among ICI-treated patients, corresponding to **Figure 2d**. Estimates with 95% confidence intervals for CTLA-4-based (± PD-(L)1) and PD-(L)1-based (without CTLA-4) regimens, with the adjusted p-value from the multivariable Cox model for each toxicity.

**Supplementary Table 24.** Full multivariable Cox proportional-hazards models for the CTLA-4 versus PD-(L)1 comparison in **Figure 2d**, reported as in **Supplementary Table 22**.

**Supplementary Table 25.** One-versus-rest multivariable Cox proportional-hazards models of cancer type–toxicity associations, corresponding to **Figure 2e**. Each cancer type is tested against all others separately for each toxicity. Hazard ratios, 95% confidence intervals and p-values are given for every term in each model, together with cohort and event counts, covariates removed by the covariate-selection rules, and fit status. Combinations with no events in the exposed group were not fitted and are marked as skipped. P-values are not corrected for multiple comparisons.

**Supplementary Table 26.** One-year cumulative incidence of systemic therapy toxicities by line of therapy, corresponding to **Figure S2e**. Estimates with 95% confidence intervals for lines 1, 2, 3 and 4 or later, with hazard ratios relative to first-line therapy, Wald and unadjusted log-rank p-values, and Benjamini–Hochberg and Bonferroni-corrected p-values. The unit of analysis is the line of therapy.

**Supplementary Table 27.** Patient-level frailty Cox proportional-hazards models of toxicity risk across lines of therapy, corresponding to **Figure S2e**. Hazard ratios, 95% confidence intervals and p-values for every model term, from a shared-frailty model with a patient-level random intercept (primary) and a marginal model with cluster-robust standard errors grouped by patient (sensitivity). The estimated frailty variance is reported for the shared-frailty models.

**Supplementary Table 28.** Time-varying Cox proportional-hazards models of the association between systemic therapy toxicity and progression-free survival, corresponding to **Figure 2f**. Each toxicity is modelled as a time-varying exposure at grade 1+ and grade 3+, stratified by cancer type. Results are given for all three treatment cohorts — first-line ICI, first-line non-ICI, and all patients — of which the ICI cohort is displayed in **Figure 2f**.

**Supplementary Table 29.** Time-varying Cox proportional-hazards models of the association between systemic therapy toxicity and overall survival, corresponding to Figure S2f. Hazard ratios with 95% confidence intervals for each toxicity at grade 1+ and grade 3+, in first-line ICI-treated, first-line non-ICI-treated, and all patients. **Figure S2f** displays the ICI cohort.

**Supplementary Table 30.** Distribution of toxicity grades around grade 3 or higher events, corresponding to **Figure S2g**. Percentage of the grade 3+ cohort at each grade, at 90-day intervals before and after the severe episode, with cohort size per toxicity. Hyperthyroidism was not analyzed because too few grade 3+ events were observed.

**Supplementary Tables 31–32**. Discrimination of the toxicity-agnostic random survival forest model, which predicts onset of any of the six adverse events, at grade 1+ (31) and grade 3+ (32). Held-out test-set Harrell’s c-index with 95% confidence intervals from 100 bootstrap resamples, reported separately for immune checkpoint inhibitor–treated and non–immune checkpoint inhibitor–treated lines of therapy, together with the number of lines of therapy and events contributing to each estimate. The grade 3+ model corresponds to **Figure 3b**.

**Supplementary Tables 33–34**. Twelve-month cumulative incidence of any of the six adverse events by random survival forest risk tertile, at grade 1+ (33) and grade 3+ (34). One row per treatment group (immune checkpoint inhibitor–treated, non–immune checkpoint inhibitor– treated) and risk stratum (low, medium, high), giving the number of lines of therapy, the number of events, and the cumulative incidence with its 95% confidence interval at 365 days. The grade 3+ estimates correspond to **Figure 3b**.

**Supplementary Table 35.** Discrimination of the adverse-event–specific random survival forest models trained on grade 1+ incidence of liver toxicity, pneumonitis, hypothyroidism, colitis and adrenal insufficiency. Held-out test-set Harrell’s c-index with 95% confidence intervals from 100 bootstrap resamples, reported separately for immune checkpoint inhibitor–treated and non– immune checkpoint inhibitor–treated lines of therapy. Hyperthyroidism was not modelled because too few events were observed. Corresponds to **Figure 3c**.

**Supplementary Table 36.** Twelve-month cumulative incidence of each adverse event by random survival forest risk tertile. One row per adverse event, treatment group (immune checkpoint inhibitor–treated, non–immune checkpoint inhibitor–treated) and risk stratum (low, medium, high), giving the number of lines of therapy, the number of events, and the cumulative incidence with its 95% confidence interval at 365 days. Corresponds to **Figure 3c**.

**Supplementary Tables 37–38**. Held-out test-set c-index of the toxicity-agnostic random survival forest model across clinical subsets, at grade 1+ (37) and grade 3+ (38). One row per clinical subset and data split, giving the subset type (overall, lot_overall, coarse_tuple or fine_tuple), cancer type, line of therapy, treatment regimen, training or test split, c-index, sample size and event count. Corresponds to **Figure S3a–S3b**.

**Supplementary Tables 39–43**. Held-out test-set c-index of the adverse-event–specific random survival forest models across clinical subsets, for liver toxicity (39), pneumonitis (40), hypothyroidism (41), colitis (42) and adrenal insufficiency (43). Columns as in Supplementary Tables 37–38. Corresponds to **Figure S3a–S3b**.

**Supplementary Table 44.** Summary of all genome-wide association analyses, by adverse event and treatment cohort. Sample size, event count, genomic control inflation factor, and counts of genome-wide significant and suggestive variants, with the covariate set and an indicator of which analyses were primary.

**Supplementary Table 45.** All variants reaching genome-wide significance (P < 5 × 10⁻⁸). SPACox and Cox P values, score statistic and variance, z, minor allele frequency, missingness, chromosome, position and nearest gene, for hypothyroidism, pneumonitis and adrenal insufficiency.

**Supplementary Table 46.** GTEx v8 cis-eQTL results for the lead variants, giving tissue, normalized effect size and P value.

**Supplementary Table 47.** Genotype composition of each lead variant across 1000 Genomes Phase 3 superpopulations, with genotype counts and alternate allele dosage.

**Supplementary Table 48.** Replication of the genome-wide significant loci in the Dana-Farber Cancer Institute PROFILE cohort. One row per lead variant, phenotype, severity threshold and ancestry, giving the best linkage-disequilibrium proxy, its r² and imputation INFO, and the DFCI hazard ratio, confidence interval and P value.

**Supplementary Table 49.** Fine–Gray subdistribution and cause-specific Cox estimates for each lead variant, fitted separately in ICI-treated, non-ICI-treated and combined cohorts with death as a competing risk.

**Supplementary Table 50.** FinnGen Data Freeze 12 phenome-wide association results for each lead variant, giving endpoint, P value, effect size and case count.

**Supplementary Table 51.** UK Biobank associations retrieved through the Open Targets Platform, giving study, contributing cohort, trait and P value.

**Supplementary Table 52.** Look-up of the three previously reported immune-related adverse event loci in MSK-Tox. One row per locus, adverse event, grade threshold and treatment cohort, giving the best proxy, its r², z, P value and direction. No hazard ratio is reported because SPACox provides a score test rather than a fitted Cox model.

**Supplementary Table 53.** Regional association statistics for adrenal insufficiency across the HLA/MHC locus. SPACox P values, score statistic and its variance, z-score, minor allele frequency, and missing rate for each tested variant, together with its chromosome, position, and annotated HLA gene/region label (or Non-MHC/Intergenic/Unknown for variants outside the named HLA genes) used to define the zoomed regional plot. Corresponds to **Figure 5a**.

**Supplementary Tables 54–56**. Position-wise omnibus association of DRB1 antigen-recognition-domain residues with adrenal insufficiency in the MSK-IMPACT cohort. For each tested IMGT mature-protein position and severity threshold (grade 1+, 2+ and 3+), the majority (baseline) residue, likelihood-ratio statistic and degrees of freedom comparing Cox models with versus without that position’s residue indicator variables, the resulting P value and Benjamini-Hochberg FDR, cohort size, and majority/minority-residue patient and event counts. Corresponds to **Figures 5b** and **5c**.

**Supplementary Table 57.** External replication of the DRB1 positional association scan in the Dana-Farber Cancer Institute cohort. For each tested HLA-DRB1 antigen-recognition-domain position, toxicity, and severity threshold (mild/moderate/severe, corresponding to grade 1+/2+/3+), the replication cohort’s test statistic and P value, with FDR computed within each locus × severity × toxicity group. Plotted alongside the MSK-IMPACT results in the lollipop plot. Corresponds to **Figure 5b**.

**Supplementary Tables 58–60**. Full multivariable Cox proportional-hazards models underlying the residue-level hazard ratios at HLA-DRB1 positions 11, 13, and 71 in the grade 1+ adrenal insufficiency cohort. Coefficient, hazard ratio, standard error, 95% confidence interval, z-statistic and p-value for every term in each position’s model: age at line-of-therapy start (in decades), sex, ancestry, cancer line-of-therapy count, therapy-exposure flags (containment of chemotherapy, biologic, targeted, hormone, CTLA-4-containing immunotherapy), and an indicator for each non-majority residue observed at that position, relative to the position’s majority (baseline) residue. Corresponds to **Figure 5e**.

**Supplementary Table 61.** DRB1 allele-level risk-residue composition at positions 11, 13 and 71. For each DRB1 allele carrying at least one risk-associated residue, its amino acid at each of the three positions, whether that residue matches the risk-associated residue at that position, the allele’s total risk score (number of risk-matching positions, 0-3). Corresponds to **Figure 5e** and **5f**.

**Supplementary Tables 62–64**. Cumulative incidence of adrenal insufficiency by DRB1*15 allele dosage among patients on a first CTLA-4-inhibitor-containing line of therapy. Aalen-Johansen cumulative-incidence estimate and 95% confidence interval at each observed event time in months since line-of-therapy start, for each dosage group (0, 1 or 2 copies of DRB1*15:01/02/03); number of patients remaining at risk at 3-month intervals from treatment start through 18 months, for each dosage group; and, per dosage group, sample size and event count, each carrier group’s pairwise log-rank P value against the non-carrier reference group, and the overall multivariate log-rank P value. Corresponds to **Figure 5g**.

**Supplementary Table 65.** Receiver operating characteristic analysis in the 300-patient test set. Per-toxicity AUC, selected operating threshold, confusion-matrix counts at that threshold, and the resulting sensitivity, specificity, precision and F1. Corresponds to **Figure S1e**.

**Supplementary Note 1.** Full text of the toxicity detection prompt submitted to Llama 4 Maverick 17B, returning a confidence score between 0 and 1 for each of the six adverse events with a free-text rationale. Includes the per-toxicity definitions, their exclusions, and the rules for scoring findings that are ruled out, hypothetical or resolved. Corresponds to the Toxicity annotation section of the **Methods**.

**Supplementary Note 2.** Full text of the CTCAE grade imputation prompt, applied to batches with a positive detection call. Presents the CTCAE v5.0 grading anchors for each toxicity and returns a grade of 1 to 5; presence is not re-adjudicated. Corresponds to the Grade and subtype annotation section of the **Methods**.

**Supplementary Note 3.** Full text of the adrenal insufficiency subtype classification prompt, applied to batches positive for adrenal insufficiency. Classifies each case as primary, secondary (central, from hypophysitis), or not attributable to systemic anti-cancer therapy. Corresponds to the Grade and subtype annotation section of the **Methods**.

## Data Availability

All data underlying the figures and reported statistics are provided in Supplementary Tables 1-65. All code necessary to reproduce the findings in the manuscript are shared at https://github.com/ziadbakouny18/msk-tox. Raw sequencing data are restricted to protect patient privacy in accordance with applicable federal and state laws and institutional policies. Additional de-identified individual-level clinical data may be made available from the corresponding authors upon reasonable request and subject to institutional review and execution of an appropriate data use agreement.

## Acknowledgements

This work was supported by the MSK Support Grant/Core Grant (P30 CA008748) and the Halvorsen Center for Computational Oncology. The authors would like to thank MSK’s Cancer Data Science Initiative (CDSI) and the patients who shared their data to make this work possible. WT is supported by the NIH/NCI (R37 CA271186, U54 CA274492, P30 CA008748), Break Through Cancer, the Fund for Innovation in Cancer Informatics, the Cancer AI Alliance, the Tow Center for Developmental Oncology, and the Maurice Campbell Initiative at Memorial Sloan Kettering Cancer Center. ZB is supported by The Fund for Innovation in Cancer Informatics, MSK’s AI Technology Development Fund, MSK’s cycle for survival grant program, the Shulamit Katzman GMTEC fellowship, and the Louis V. Gerstner, Jr. Physician Scholars program. ER is supported by a Pershing Square Cancer Research Prize. This work utilized resources from the High-Performance Computing Group at Memorial Sloan Kettering Cancer Center. RRK is supported, in part, by a DoD KCRP-ECI Award (W81XWH-21-0942), AACR-KidneyCAN Award (26-80-81), and Kidney Cancer Association (25-PED-001).

## Competing financial interests

Z.B., X.A.G., F.H., S.M., W.T., J.C.Z, and E.R. filed a patent that pertains to the annotation and prediction of systemic therapy toxicities, which is related to the scope of this manuscript. Z.B. reports honoraria from UpToDate and the Fund for Innovation in Cancer Informatics unrelated to the current study. K.C.A. has served as a paid consultant to Bristol Myers Squibb, Genentech, Verastem, Merck, Revolution Medicines, Nuvalent, and Tango Therapeutics. She has received research support (to her institution for conduct related to clinical trials) from Bristol Myers Squibb, Treeline Biosciences, Gilead, Revolution Medicines, Mirati, Verastem, and BridgeBio Pharma unrelated to the current study. R.N. reports consulting for Seres Therapeutics and I-Mab Biopharma unrelated to the current study. A.J.S. reports consulting/advising role to J&J, mBRACE therapeutics, Revolution Medicine (IDMC), Bayer, KSQ therapeutics, BioNtech, BMS, Merck, Astrazeneca, Oxford Biotherapeutics, Roche, Synthekine, cTRL therapeutics, Regeneron, Enara Bio, Perceptive Advisors, Foresight Diagnostics, Oppenheimer and Co, Umoja Biopharma, Legend Biotech, Iovance Biotherapeutics, Obsidian Therapeutics, Prelude Therapeutics, Immunocore, Lyell Immunopharma, Amgen, Boehringer Ingelheim and Heat Biologics. Research funding: GSK (Inst), Obsidian (Inst), Lilly (Inst), Astrazeneca (Inst), BioNtech (Inst), Clasp Therapeutics (Inst), PACT pharma (Inst), Iovance Biotherapeutics (Inst), Achilles therapeutics (Inst), Merck (Inst), Synthekine (Inst), BMS (Inst), Harpoon Therapeutics (Inst), AffiniT therapeutics (Inst), Fortvita (Inst) Legend Therapeutics (Inst), Synthekine (Inst) and Amgen (Inst) unrelated to the current study. C.B.T. is a member of the board of directors and a shareholder of Regeneron and Charles River Laboratories, and a founder of Agios Pharmaceuticals unrelated to the current study. D.M.F. served as a consultant for AstraZeneca and Seres and has research support from Janssen unrelated to the current study. M.F.B reports Personal fees from AstraZeneca and Intellectual property rights for SOPHiA Genetics unrelated to the current study. R.J.M. reports paid Consulting from Merck and AstraZeneca; Clinical trial support to MSKCC from Merck, Bristol Myers Squibb, Exelixis, Eisai unrelated to the current study. RRK has provided consulting or advisory roles for Eisai and Merck, and has received institutional research funding from Pfizer, Takeda, Novartis, Exelixis, Xencor, Arsenal Bio, and Allogene Therapeutics unrelated to the current study.

---

**Supplementary Figure 1:**
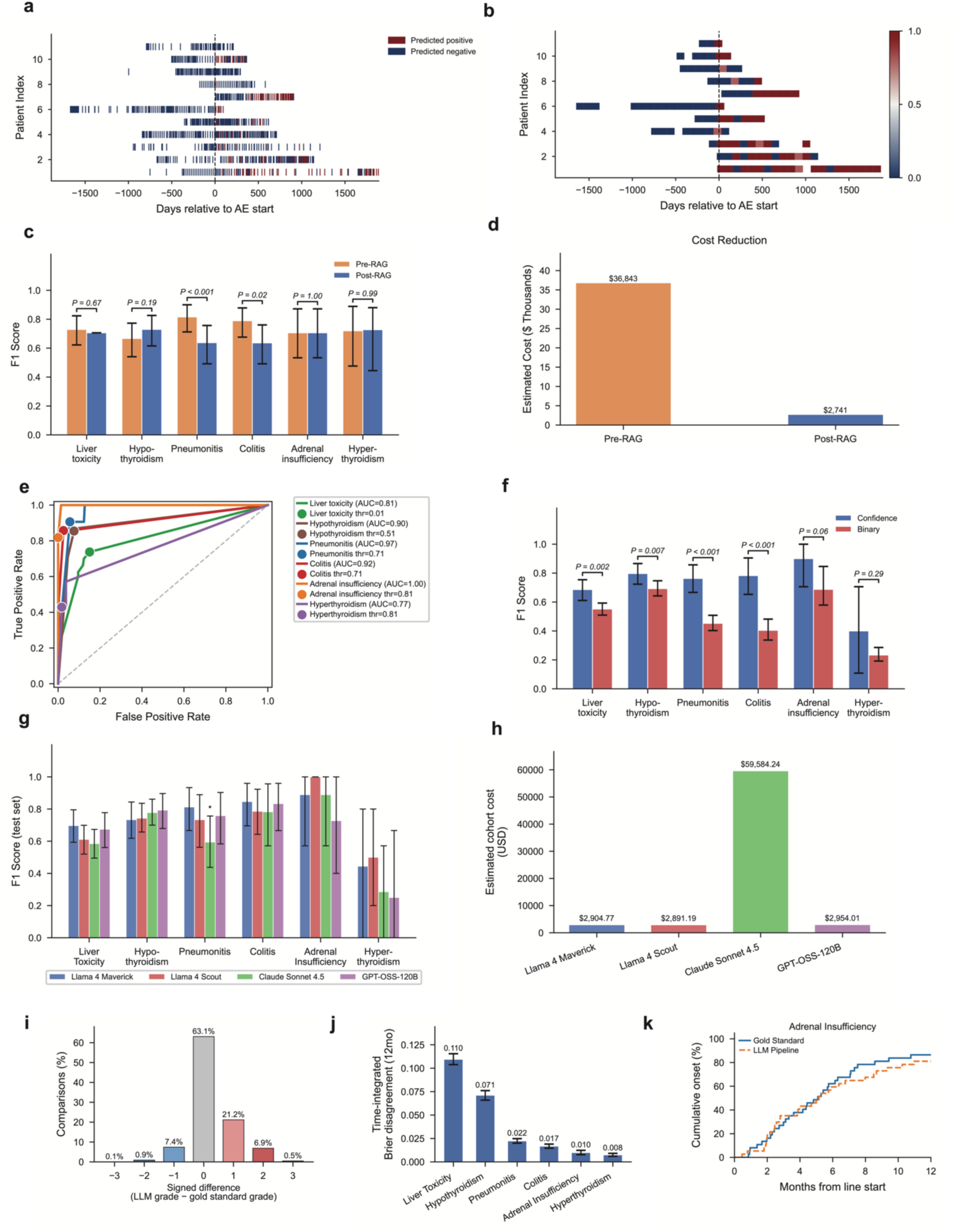
Validation of the systemic therapy toxicity annotation pipeline. **a**, **b**, Representative examples of 11 patients who had developed adrenal insufficiency per the gold standard. A note-level analysis (**a**, each row representing one patient, each data point representing one clinical note) using Llama 4 Maverick. The vertical dotted line represents the time at which the patients had developed adrenal insufficiency based on the gold standard. The same data for the same 11 patients is represented post-RAG in panel **b**. **c**, Comparison of F1 scores pre-and post-RAG in a subset of 110 patients. 95% confidence intervals for the F1 scores and the associated P-values (two-sided paired bootstrap test) were derived by drawing 2,000 stratified bootstrap resamples. **d**, Estimated inference costs pre-and post-RAG, extrapolated to 55,406 patients. **e**, Receiver operating characteristic (ROC) curves for detection of individual adverse events in the 300-patient test set. Area under the ROC curve (AUC) is shown for each adverse event. Points indicate operating performance at the corresponding thresholds optimized on the training set. **f**, Comparison of F1 scores between confidence-based and binary LLM outputs across individual immune-related adverse events in the 300-patient test set. Error bars indicate 95% confidence intervals derived from 2,000 stratified bootstrap resamples. *P* values were calculated using two-sided paired bootstrap tests with 2,000 stratified resamples. **g**, Comparison of test-set F1 scores across Llama 4 Maverick, Llama 4 Scout, Claude Sonnet 4.5 and GPT-OSS-120B for individual adverse events. Model-specific thresholds were determined in the training set and performance was evaluated in the held-out test set. Error bars indicate 95% confidence intervals derived from 2,000 stratified bootstrap resamples. P values were calculated using two-sided paired bootstrap tests comparing each model with Llama 4 Maverick. Asterisk denotes P < 0.05. **h**, Estimated LLM inference costs for the full patient cohort across Llama 4 Maverick, Llama 4 Scout, Claude Sonnet 4.5 and GPT-OSS-120B. **i**, Distribution of signed differences between LLM-assigned and gold-standard adverse event grades across 1,454 matched patient–toxicity comparisons. Signed differences were calculated as LLM grade minus gold-standard grade; a value of zero indicates exact agreement. **j**, Time-dependent validation of LLM-predicted adverse events using the time-integrated Brier score over 12 months from the start of each line of therapy. Error bars indicate 95% confidence intervals derived from 500 bootstrap resamples. **k**, Cumulative incidence comparison of time to adverse event onset according to the gold standard and LLM predictions for adrenal insufficiency among concordant events

**Supplementary Figure 2:**
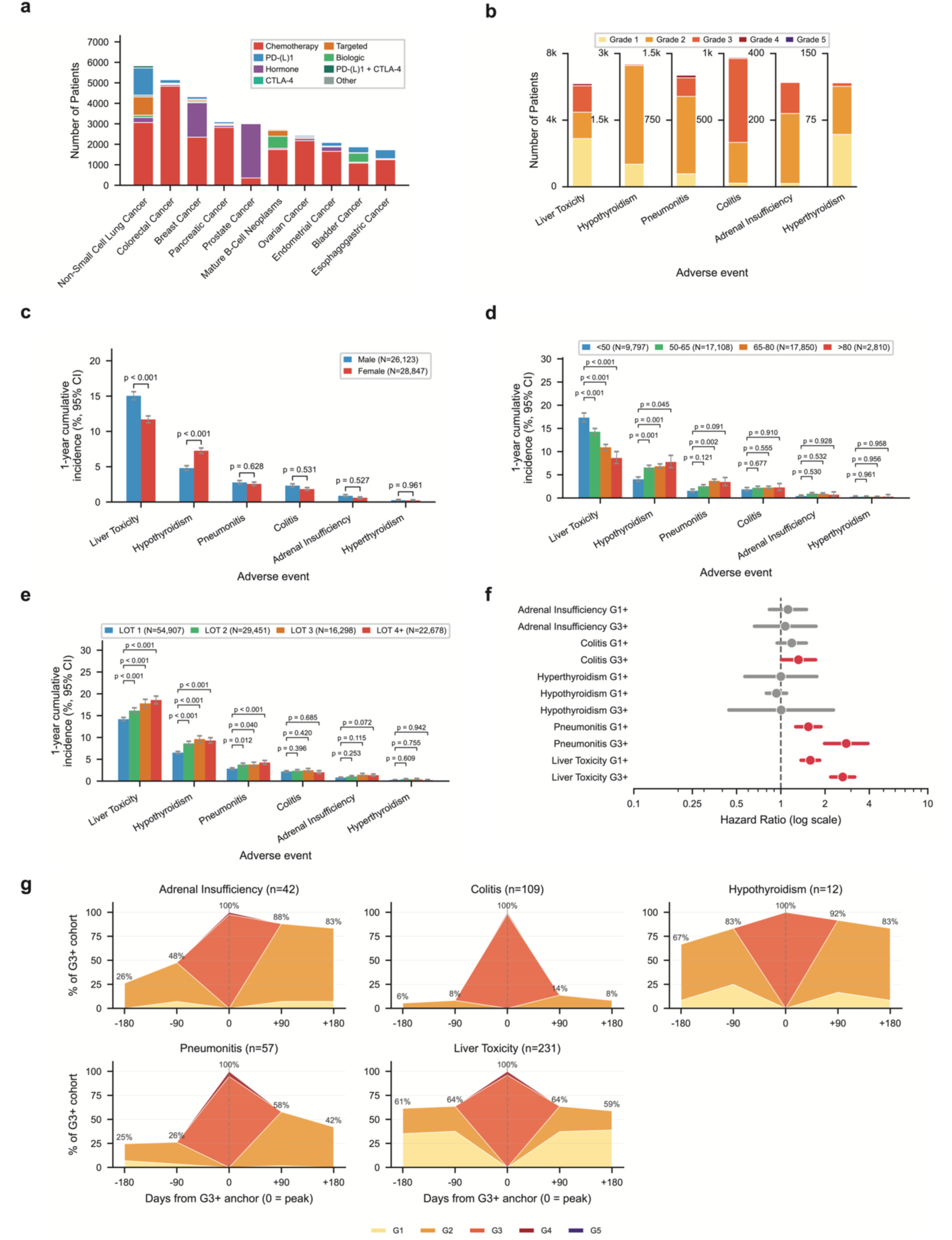
Associations of systemic therapy toxicities with clinical and demographic features as well as clinical outcomes. **a**, Distribution of first-line systemic therapy classes across the 10 most common cancer types in MSK-Tox. Stacked bars indicate the number of patients receiving each systemic therapy class. **b**, Distribution of CTCAE grades for adverse events occurring during the first line of therapy in MSK-Tox. Stacked bars indicate the number of patients with grade 1–5 events for each toxicity type. **c**, One-year cumulative incidence of adverse events during the first line of therapy, stratified by sex. Error bars indicate 95% confidence intervals. P values are from multivariable Cox proportional-hazards models comparing females with males for each toxicity. **d**, One-year cumulative incidence of adverse events during the first line of therapy, stratified by age at the start of first-line therapy. Error bars indicate 95% confidence intervals. P values are from multivariable Cox proportional-hazards models comparing each age group with patients aged <50 years (reference) for each toxicity. **e**, One-year cumulative incidence of adverse events stratified by line of therapy (LOT). Error bars indicate 95% confidence intervals. P values are from multivariable Cox proportional-hazards models comparing LOT 1 (reference), LOT 2 and LOT 3 with LOT ≥4 for each toxicity. **f**, Associations between immune-related adverse events and overall survival among patients receiving immune checkpoint inhibitors. Forest plots show hazard ratios and 95% confidence intervals from time-varying Cox proportional-hazards models. Bars in red indicate P < 0.05. **g**, Temporal distribution of adverse event grades around grade ≥3 events. Time zero represents the peak grade ≥3 event, and bars indicate the distribution of toxicity grades over time relative to this event.

**Supplementary Figure 3:**
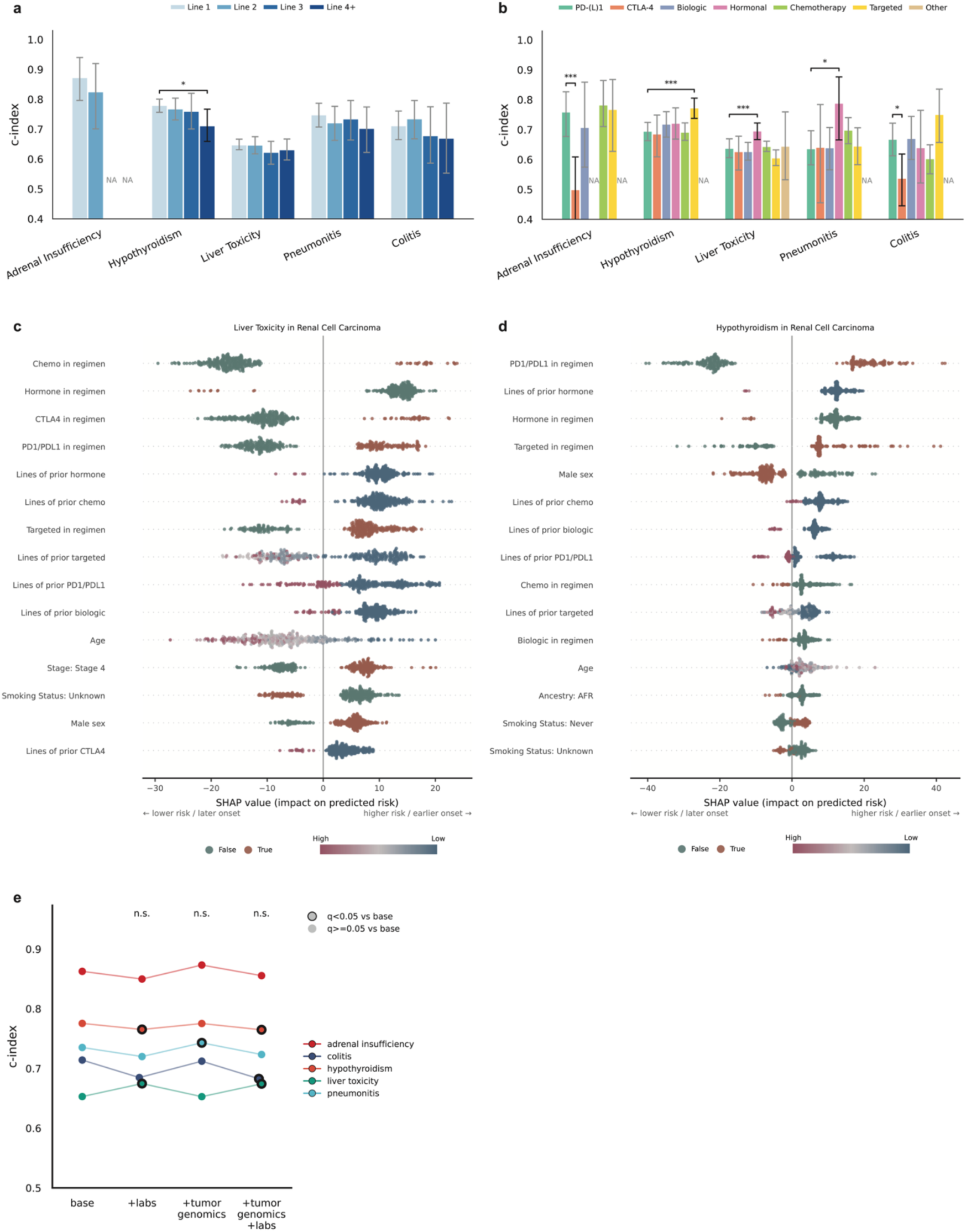
Validation of the random survival forest models across clinical settings and explainability of the models. **a,** Performance of risk models across lines of therapy. **b,** Performance of risk models across treatment modalities contained in lines of therapy. Each setting contains at least 25 patients under treatment. Model performance across settings were compared against first displayed setting (line 1; PD-(L)1) for difference in c-index, using 500 of nonparametric bootstrap to derive p-values and a significance cutoff of p<0.05. Significant comparisons were annotated with p-values and displayed with black confidence internals. **c,** Summary of SHAP values for the liver toxicity model in renal cell carcinoma patients. **d,** Summary of SHAP values for the hypothyroidism model in renal cell carcinoma patients. **e.** Performance comparison between models trained with the additional of lab values, tumor genomics, or both sets of features. C-indices were compared against the base model of each adverse event with a significance threshold 0.05 for BH-corrected q-value. Models with significance difference to the base model was circled in black. All risk models were trained separately on grade 1+ incidence of each adverse event. Harrell’s C-index was calculated on held out test set, and 95% confidence interval was derived from 500 rounds of sample bootstrap.

**Supplementary Figure 4:**
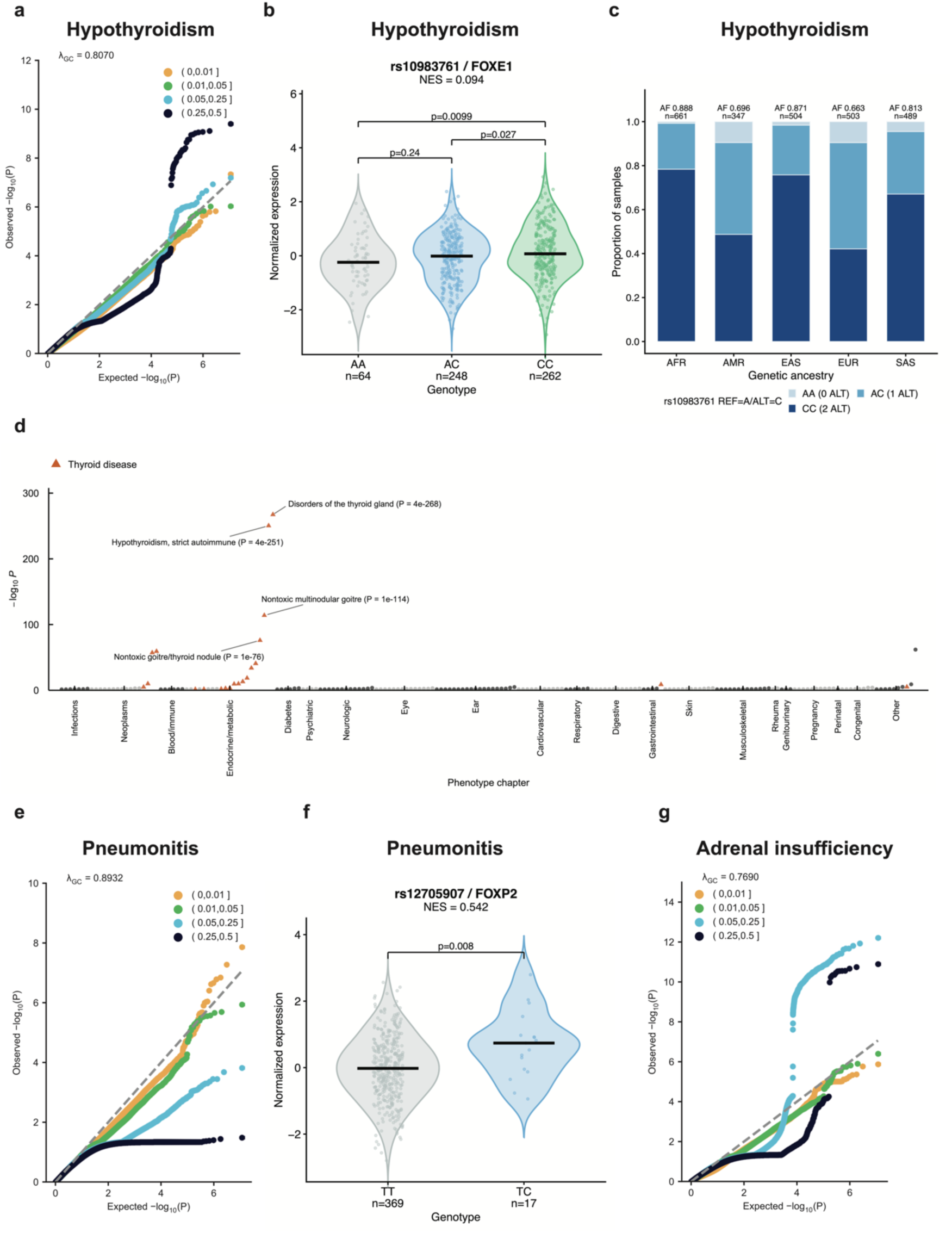
Analytical validity of genome-wide association studies, population frequencies of lead variants, and external validation. **a**, Quantile–quantile plot of genome-wide association results for hypothyroidism in the combined treatment cohort (35,669 patients, 1,765 events). Variants are colored by minor allele frequency bin; the dashed grey line is the null expectation and the genomic control inflation factor (λGC) is annotated. **b**, GTEx v8 cis-eQTL for the hypothyroidism lead variant rs10983761 on *FOXE1* expression in thyroid (normalized effect size, NES = 0.094). Violins show the distribution of normalized expression by genotype with individual donors overlaid; black bars denote medians and sample sizes are given per genotype. Pairwise genotype comparisons were made with two-sided Wilcoxon rank-sum tests and annotated with p-values. **c**, Genotype composition of rs10983761 across 1000 Genomes Phase 3 superpopulations, with the alternate allele frequency and number of samples shown above each ancestry. **d**, FinnGen R12 phenome-wide association results for rs10983761, with endpoints ordered by ICD chapter and plotted as −log10(P-value). Thyroid endpoints are highlighted; the most significant thyroid endpoints are labelled with their p-values. **e**, Quantile–quantile plot for pneumonitis in the immune checkpoint inhibitor cohort (9,723 patients, 661 events), displayed as in a. **f**, GTEx v8 cis-eQTL for the pneumonitis lead variant rs12705907 on *FOXP2* expression in heart left ventricle (NES = 0.542), displayed as in b. **g**, Quantile–quantile plot for adrenal insufficiency in the immune checkpoint inhibitor cohort (9,760 patients, 258 events), displayed as in a. All genome-wide association analyses were run separately for each adverse event on grade 1+ incidence, modelling time to event onset with SPACox and adjusting for sex, age, line of therapy, treatment class, cancer type, sequencing panel and the first ten genetic principal components, with genome-wide significance defined as P < 5 × 10⁻⁸.

**Supplementary Figure 5:**
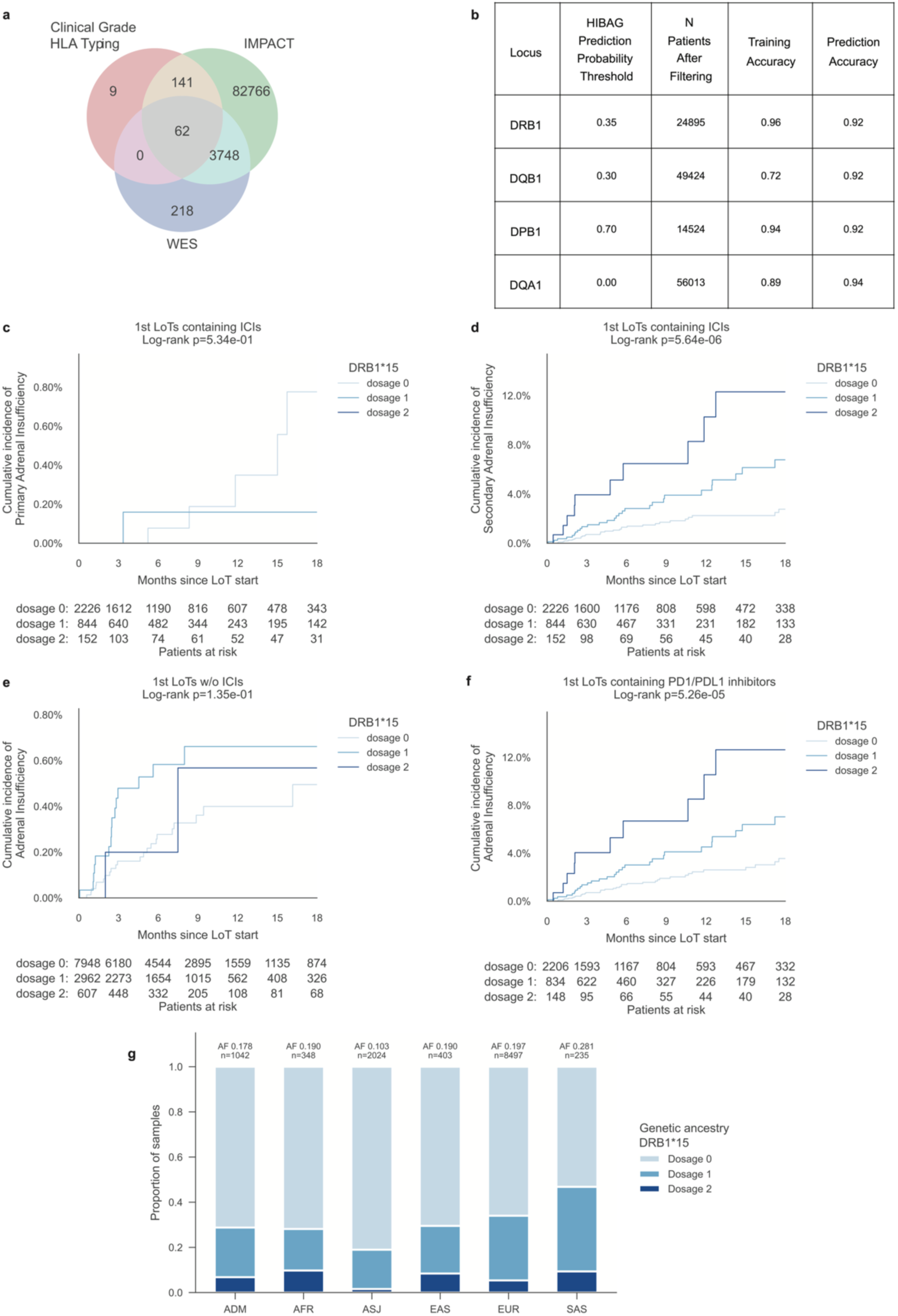
HLA typing imputation quality control and sensitivity analyses of the association between DRB1*15 and adrenal insufficiency. **a,** Overlap of patients with HLA genotypes obtained from clinical-grade HLA typing, MSK-IMPACT targeted sequencing and whole-exome sequencing. **b,** HIBAG posterior prediction-probability thresholds selected for each class II HLA locus, together with the resulting number of patients retained and the corresponding training and prediction accuracies. **c,d,** Cumulative incidence of primary **(c)** and secondary **(d)** adrenal insufficiency according to DRB1*15 dosage among first lines of therapy containing an ICI. **e,** Cumulative incidence of adrenal insufficiency according to DRB1*15 dosage among first lines of therapy not containing an immune checkpoint inhibitor. **f,** Cumulative incidence of adrenal insufficiency according to DRB1*15 dosage among first lines of therapy containing PD-1 or PD-L1 inhibitor. **g,** Distribution of DRB1*15 dosage across genetically inferred ancestry groups in MSK cohort. Allele frequency and sample size are indicated above each bar. ADM, admixed ancestry; AFR, African ancestry; ASJ, Ashkenazi Jewish ancestry; EAS, East Asian ancestry; EUR, European ancestry; SAS, South Asian ancestry; HIBAG, HLA imputation using attribute bagging; ICI, immune checkpoint inhibitor; LoT, line of therapy; WES, whole-exome sequencing.

## Notes

### Author Declarations

The Institutional Review Board of Memorial Sloan Kettering Cancer Center gave ethical approval for this work.

