## Supplementary Note 1 for "Population-Scale Precision Safety in Oncology Reveals Clinical and Genetic Determinants of Systemic Therapy Toxicity"

Indicate the probability that the following toxicities are present for the patient in the clinical note.
The probability should be a continuous number between 0 and 1.
CRITICAL INSTRUCTIONS:
1. Only assign a probability > 0 if there is CLEAR EVIDENCE of the toxicity being present at the time the note was written
2. If the toxicity is mentioned but explicitly ruled out, assign probability = 0
3. If the toxicity is only mentioned as a differential diagnosis or potential risk (but not confirmed), assign probability = 0
4. If the toxicity is mentioned as resolved or historical (not current), assign probability = 0
5. If the toxicity is present but clearly unrelated to systemic anti-cancer therapy, assign probability = 0
6. Imaging findings should be specific and not easily explained by other causes
7. It is possible for all 6 toxicities to be absent from the note
8. Be especially cautious with common symptoms like fatigue, which can have many causes
9. If a toxicity is mentioned as "resolved," "improved," or "treated," assign probability = 0
10. If the adverse event is related to prior surgical resection (i.e. thyroidectomy) and not systemic anti-cancer therapy, assign probability = 0
11. Be conservative - when in doubt, assign a lower probability
The toxicities are:
1. Adrenal insufficiency
2. colitis
3. hyperthyroidism
4. hypothyroidism
5. pneumonitis
6. Liver toxicity
These are the descriptions for the 6 toxicities:
Adrenal Insufficiency:
Adrenal insufficiency is a state of reduced adrenal function or cortisol levels related to systemic anti-cancer therapies. This may or may not be associated with hypophysitis, which would be associated with other abnormal hormone levels (such as low ACTH). Steroids or hormone replacement therapy can be used as treatment.
EXCLUDE: primary adrenal diseases unrelated to cancer therapy such as metastases to the adrenal glands leading to adrenal insufficiency or secondary adrenal insufficiency related to prolonged steroid treatment, unless specified as exacerbated by systemic anti-cancer therapies.
Colitis:
Colitis reflects inflammation of the colon that is related to systemic anti-cancer therapies. Colitis often requires treatment with immunosuppressive medications, such as corticosteroids. More severe colitis would typically lead to a change of treatment and/or hospitalization.
EXCLUDE: infectious colitis or other causes of colon inflammation that are not related to systemic anti-cancer therapies (such as pre-existing ulcerative colitis or Crohn's disease), unless specified as exacerbated by systemic anti-cancer therapies.
Hyperthyroidism:
Hyperthyroidism represents an overactive thyroid linked to systemic anti-cancer therapies. Lab findings would include low TSH (thyroid-stimulating hormone) and elevated free T4 or T3. More severe hyperthyroidism may require interventions, such as antithyroid drugs or beta-blockers. When hyperthyroidism is related to immune checkpoint inhibitors, it often reflects thyroiditis and is followed by a period of hypothyroidism (which is a separate adverse event).
EXCLUDE: preexisting hyperthyroidism unrelated to therapy, unless specified as exacerbated by radiation therapy or systemic anti-cancer therapies.
Hypothyroidism:

Hypothyroidism represents underactive thyroid associated with systemic anti-cancer therapies. Lab findings would include elevated TSH and low free T4. Hypothyroidism often requires treatment with thyroid hormone replacement therapy as treatment. When hypothyroidism is related to immune checkpoint inhibitors, it often reflects thyroiditis and sometimes follows a period of hyperthyroidism (which is a separate adverse event).

EXCLUDE: pre-existing hypothyroidism unrelated to therapy, such as those related to cancer invading the thyroid, thyroidectomy, or pre-existing disorders (like Hashimoto's disease), unless specified as exacerbated by systemic anti-cancer therapies.

Pneumonitis:

Pneumonitis is an inflammatory process of the lungs caused by systemic anti-cancer therapies. More severe pneumonitis would typically lead to a change of treatment and/or hospitalization.

EXCLUDE: infectious pneumonia and other instances of pneumonitis or lung inflammation that are not related to systemic anti-cancer therapies, unless specified as exacerbated by anti-cancer therapies.

Liver Toxicity:

Liver toxicity represents liver dysfunction or injury linked to systemic anti-cancer therapies. It is almost always reflected by lab findings like elevated liver enzymes (alanine aminotransferase or ALT, aspartate aminotransferase AST), bilirubin (total, direct, or indirect), alkaline phosphatase (or alk phos), or gamma-glutamyl transferase (GGT). Transaminitis is sometimes a term used to refer to liver toxicity. More severe liver toxicity often requires immunosuppressive therapy (such as steroids), dose adjustments in response to liver toxicity, pauses in therapy, or even discontinuation of therapy. Very severe liver toxicity can lead to hospitalization or a change in anti-cancer therapy.

EXCLUDE: other causes of liver dysfunction, such as preexisting liver diseases, liver dysfunction related to infectious causes, or metastases to the liver unless specified as exacerbated by systemic anti-cancer therapies.

Rationale:

Provide a clear rationale for why each toxicity is present (Probability > 0) or not present (Probability = 0). Focus on the specific evidence in the note that supports or refutes the presence of each toxicity.

Format the output as a JSON object with fields for each toxicity and a single rationale field. Always include all six toxicity fields and the rationale in the JSON output. Make sure that the JSON has the actual values for the predicted probabilities in it. There is never a reason to display text outside of the JSON.

The JSON should have this format:

```
{
"adrenal insufficiency": 0.0,
"colitis": 0.0,
"hyperthyroidism": 0.0,
"hypothyroidism": 0.0,
"pneumonitis": 0.0,
"liver toxicity": 0.0,
"rationale": "Your detailed rationale here
```
