## Supplementary Note 2 for "Population-Scale Precision Safety in Oncology Reveals Clinical and Genetic Determinants of Systemic Therapy Toxicity"

CRITICAL INSTRUCTION: You are given batched clinical notes for a single patient plus binary indicators (1/0) from a prior run for each toxicity below.

**IMPORTANT RULES:**
1. If a toxicity = 1, you MUST assign AT LEAST grade 1 for that toxicity
2. Do NOT re-adjudicate presence?trust the 1/0 inputs absolutely
3. If toxicity = 1 but you find no evidence in the text, assign grade 1 (mild/subclinical). DO NOT ASSIGN 0 FOR THIS CASE.
4. Only use the text to determine severity (grade 1-5), NOT presence. If the binary indicators indicate presense then believe it as ground truth.

*Grading Rules:*
- If toxicity = 0 ? grade = 0
- If toxicity = 1 and no evidence found ? grade = 1 (mild/subclinical)
- If toxicity = 1 and evidence found ? assign worst grade 1-5 based on CTCAE criteria

Toxicities to grade when flagged = 1:
- Adrenal insufficiency
- Colitis
- Hyperthyroidism
- Hypothyroidism
- Pneumonitis
- Liver toxicity (use worst among: AST increased, ALT increased, ALP increased, GGT increased, Blood bilirubin increased, Hepatic failure, Hepatic pain)

CTCAE v5.0 grading anchors
Adrenal insufficiency
G1: Asymptomatic; labs only; no treatment.
G2: Symptomatic; hormone replacement indicated.
G3: Severe; hospitalization indicated.
G4: Life-threatening; urgent intervention.
G5: Death.
Colitis
G1: Asymptomatic; diagnostic findings only.
G2: Abd pain and/or ?stool frequency or blood/mucus; outpatient therapy (e.g., steroids).
G3: Severe pain/dehydration or hospitalization.
G4: Life-threatening (e.g., perforation, toxic megacolon).
G5: Death.
Hyperthyroidism
G1: Labs only (?TSH with ?FT4/T3); asymptomatic.
G2: Symptomatic; needs beta-blocker ± antithyroid.
G3: Severe; hospitalization.
G4: Thyroid storm/life-threatening.
G5: Death.
Hypothyroidism
G1: Labs only (?TSH ± ?FT4).
G2: Symptomatic; requires thyroid replacement.
G3: Severe; hospitalization.
G4: Myxedema coma/life-threatening.
G5: Death.
Pneumonitis
G1: Asymptomatic; radiographic only.
G2: Symptomatic; medical intervention; no oxygen.
G3: Oxygen required; limits self-care ADLs.
G4: Life-threatening respiratory compromise (e.g., ventilatory support).
G5: Death.

Liver toxicity (report worst grade across parameters)
AST increased / ALT increased (same thresholds)
If baseline normal: G1 > ULN-3x ULN; G2 >3-5x; G3 >5-20x; G4 >20x
If baseline abnormal: G1 >1.5-3x baseline; G2 >3-5x; G3 >5-20x; G4 >20x
ALP increased / GGT increased (same thresholds)
If baseline normal: G1 > ULN-2.5x ULN; G2 >2.5-5x; G3 >5-20x; G4 >20x
If baseline abnormal: G1 >2.0-2.5x baseline; G2 >2.5-5x; G3 >5-20x; G4 >20x
Blood bilirubin increased
If baseline normal: G1 > ULN-1.5x ULN; G2 >1.5-3x; G3 >3-10x; G4 >10x
If baseline abnormal: G1 >1-1.5x baseline; G2 >1.5-3x; G3 >3-10x; G4 >10x
Hepatic failure: G3 (clinical hepatic failure; hospitalization), G4 (life-threatening), G5 (death).
Hepatic pain: G1 (mild), G2 (moderate; limits instrumental ADLs), G3 (severe; limits self-care ADLs / hospitalization). Grades 4?5 not defined in CTCAE v5.0?do not assign above G3.
Input format:

```
{
  "text": "Full concatenated clinical notes for the patient",
  "adrenal insufficiency": 0 or 1,
  "colitis": 0 or 1,
  "hyperthyroidism": 0 or 1,
  "hypothyroidism": 0 or 1,
  "pneumonitis": 0 or 1,
  "liver toxicity": 0 or 1
}
```

Output format (JSON only; numeric grades + single shared rationale)
Return numeric grades for all six toxicities (0-5). Include one "rationale" string that concisely summarizes the evidence used to assign grades for each toxicity with grade ?1.{

```
  "adrenal insufficiency": 0,
  "colitis": 3,
  "hyperthyroidism": 1,
  "hypothyroidism": 0,
  "pneumonitis": 2,
  "liver toxicity": 3,
  "rationale": "Colitis graded 3 due to severe diarrhea with blood and hospitalization;
    Pneumonitis graded 2 based on cough + CT GGO and outpatient steroids without oxygen;
    Hyperthyroidism graded 1 for low TSH with mildly elevated FT4 and no therapy; Liver
    toxicity graded 3 with ALT 320 (~8× ULN) and AST 280, therapy held and steroids
    started."
}"""
```
