## Supplementary Note 3 for "Population-Scale Precision Safety in Oncology Reveals Clinical and Genetic Determinants of Systemic Therapy Toxicity"

Classify the type of adrenal insufficiency present in the clinical note into one of three categories:
1. PRIMARY ADRENAL (Adrenalitis)
2. SECONDARY ADRENAL (Hypophysitis/Hypopituitarism)
3. NOT THERAPY-RELATED

CRITICAL REQUIREMENT:
Only classify as PRIMARY ADRENAL or SECONDARY ADRENAL if the adrenal insufficiency is related to SYSTEMIC ANTI-CANCER THERAPY (such as immune checkpoint inhibitors, chemotherapy, or targeted therapies).

Classify as NOT THERAPY-RELATED if the adrenal insufficiency is:
- Caused by exogenous steroid treatment (even if steroids were given to treat other immune-related adverse events)
- Due to prolonged steroid use leading to HPA axis suppression
- Related to other causes unrelated to systemic anti-cancer therapy (e.g., metastases to adrenal or pituitary, surgical resection, pre-existing conditions)

DEFINITIONS:

Primary Adrenal Insufficiency (Adrenalitis):
Direct inflammation or damage to the adrenal glands themselves caused by systemic anti-cancer therapy. The adrenal glands are unable to produce adequate cortisol and aldosterone.
Key indicators include:
- Direct mention of adrenal inflammation, adrenalitis, or adrenal immune-related adverse events
- Mineralocorticoid deficiency (low aldosterone)
- Hyperkalemia (elevated potassium)
- Adrenal imaging showing enlarged, inflamed, or damaged adrenal glands
- Low cortisol with HIGH or NORMAL ACTH (adrenals not responding to ACTH)
- May see hyperpigmentation
- Typically does NOT have other pituitary hormone abnormalities

Secondary Adrenal Insufficiency (Hypophysitis/Hypopituitarism):
Adrenal insufficiency caused by pituitary gland dysfunction (hypophysitis or hypopituitarism) leading to insufficient ACTH production, which in turn causes the adrenal glands to produce inadequate cortisol. The adrenal glands themselves are intact but understimulated.
Key indicators include:
- Explicit mention of hypophysitis, hypopituitarism, or pituitary dysfunction
- Low or low-normal ACTH levels
- Low cortisol
- Other pituitary hormone deficiencies (low TSH, LH, FSH, GH, prolactin)
- Pituitary imaging abnormalities (enlarged pituitary, pituitary enhancement)
- Headaches or visual disturbances (signs of pituitary inflammation)
- Normal mineralocorticoid function (aldosterone typically preserved)
- Normal potassium levels

CLASSIFICATION INSTRUCTIONS:
1. FIRST determine if the adrenal insufficiency is related to systemic anti-cancer therapy or another cause (steroids, metastases, surgery, etc.)
   - If clearly NOT therapy-related ? Classify as NOT THERAPY-RELATED
   - If therapy-related ? Proceed to step 2
2. Read the clinical note carefully for evidence of the mechanism of adrenal insufficiency

3. Look for explicit mentions of hypophysitis, hypopituitarism, or pituitary dysfunction ? Classify as SECONDARY ADRENAL
4. Look for explicit mentions of adrenalitis or direct adrenal damage ? Classify as PRIMARY ADRENAL
5. Examine laboratory values:
   - Low ACTH + low cortisol ? SECONDARY ADRENAL
   - High/normal ACTH + low cortisol ? PRIMARY ADRENAL
   - Multiple pituitary hormone deficiencies ? SECONDARY ADRENAL
   - Hyperkalemia or low aldosterone ? PRIMARY ADRENAL
6. Consider imaging findings:
   - Pituitary abnormalities (enlargement, enhancement) ? SECONDARY ADRENAL
   - Adrenal abnormalities (enlargement, inflammation) ? PRIMARY ADRENAL
7. If both mechanisms are mentioned, classify based on the primary mechanism described
8. YOU MUST classify into one of the three categories - no other options are allowed
9. Assign a confidence score between 0 and 1:
   - 0.9-1.0: Explicit mention of the mechanism or clear laboratory/imaging evidence
   - 0.7-0.89: Strong supporting evidence but not explicitly stated
   - 0.5-0.69: Moderate evidence, some uncertainty remains
   - 0.3-0.49: Weak evidence, classification is uncertain
   - 0.0-0.29: Very uncertain, insufficient information to classify confidently, but best guess provided

Output the result as a JSON object with the following fields:

```
{
    "adrenal_insufficiency_type": "PRIMARY ADRENAL" or "SECONDARY ADRENAL" or "NOT
    THERAPY-RELATED",
    "confidence": 0.0-1.0,
    "rationale": "Detailed explanation of the classification decision, citing specific
    evidence from the note"
}
```

EXAMPLES:

Example 1 - PRIMARY ADRENAL:

Expected Output:

```
{
    "adrenal_insufficiency_type": "PRIMARY ADRENAL",
    "confidence": 0.98,
    "rationale": "Definitive evidence of primary adrenal insufficiency (adrenalitis)
    caused by systemic anti-cancer therapy. Key findings include: (1) Markedly elevated
    ACTH (245 pg/mL) with very low cortisol (<1 mcg/dL), indicating the pituitary is
    appropriately stimulating the adrenals but they are not responding; (2) Evidence of
    mineralocorticoid deficiency with low aldosterone (<2 ng/dL), hyperkalemia (5.8
    mEq/L), and hyponatremia (128 mEq/L) - these findings are specific to primary
    adrenal failure; (3) CT imaging shows bilaterally enlarged adrenal glands with
    inflammatory changes, consistent with adrenalitis; (4) Normal pituitary function
    evidenced by normal TSH, LH, and FSH; (5) Explicit diagnosis of 'immune-related
    adrenalitis' documented in the note. No evidence of hypophysitis or pituitary
    dysfunction."
}
```

Example 2 - SECONDARY ADRENAL:

Expected Output:

```
{
    "adrenal_insufficiency_type": "SECONDARY ADRENAL",
    "confidence": 0.95,
```

```
    "rationale": "Clear evidence of secondary adrenal insufficiency due to hypophysitis
    caused by systemic anti-cancer therapy. Multiple indicators support this
    classification: (1) Low ACTH (8 pg/mL) with low cortisol (2.1 mcg/dL) indicates
    pituitary dysfunction rather than primary adrenal failure; (2) Presence of multiple
    pituitary hormone deficiencies including low TSH and low free T4, suggesting
    panhypopituitarism; (3) MRI findings explicitly describe enlarged pituitary with
    enhancement consistent with hypophysitis; (4) Clinical context of immune checkpoint
    inhibitor therapy (pembrolizumab) which is a well-known cause of hypophysitis. No
    evidence of primary adrenal pathology or mineralocorticoid deficiency."
}
```

Example 3 - NOT THERAPY-RELATED:
Expected Output:

```
{
    "adrenal_insufficiency_type": "NOT THERAPY-RELATED",
    "confidence": 0.92,
    "rationale": "Adrenal insufficiency is present but is NOT related to systemic
    anti-cancer therapy. The note explicitly states that the patient has been on
    high-dose prednisone (60mg daily) for the past 3 months to treat immune-related
    colitis. The low morning cortisol (2.5 mcg/dL) with suppressed ACTH (3 pg/mL) is
    consistent with HPA axis suppression due to prolonged exogenous steroid use rather
    than immune-related hypophysitis or adrenalitis. There is no mention of pituitary
    imaging abnormalities, other pituitary hormone deficiencies, or adrenal gland
    inflammation that would suggest therapy-related primary or secondary adrenal
    insufficiency."
}
```
